# Cellular and molecular dysregulation of the esophageal epithelium in systemic sclerosis

**DOI:** 10.1101/2024.06.05.24308452

**Authors:** Matthew Dapas, Margarette H. Clevenger, Hadijat M. Makinde, Tyler Therron, Dustin A. Carlson, Mary Carns, Kathleen Aren, Cenfu Wei, Lutfiyya N. Muhammad, Carrie L. Richardson, Parambir S. Dulai, Monique Hinchcliff, John E. Pandolfino, Harris R. Perlman, Deborah R. Winter, Marie-Pier Tetreault

**Author notes:** These authors jointly supervised the work (Harris R. Perlman, Deborah R. Winter, Marie-Pier Tetreault).

## Abstract

Systemic sclerosis (SSc) is a rare autoimmune disease characterized by vasculopathy and fibrosis of the skin and internal organs. Individuals with SSc often suffer from chronic acid reflux and dysphagia due to loss of esophageal motility. However, the pathogenesis of esophageal dysmotility in SSc is poorly understood. To determine whether distinct changes in esophageal epithelial cells contribute to impaired motility in SSc, we investigated the stratified squamous esophageal epithelium from proximal and distal biopsies using single-cell RNA sequencing (n=306,372 cells) in individuals with SSc compared those with gastroesophageal reflux disease (GERD) as well as healthy controls. The proportion of epithelial cells in the apical, superficial compartment of the esophageal epithelium was significantly reduced in SSc (9.4% vs 21.6% in HCs). Differential gene expression in SSc was primarily limited to the superficial compartment (3,572 genes vs. 232 in all other compartments, based on pseudobulk analysis), with significant upregulation of extracellular matrix and keratinization genes. These cellular and molecular changes in SSc were highly correlated with those seen in GERD, indicating they were secondary to reflux; however, their magnitudes were more pronounced in the proximal esophagus, suggesting that esophageal dysmotility leads to greater proximal acid exposure, which may contribute to aspiration. SSc-specific gene dysregulation implicated immunoregulatory pathways likely pertinent to pathogenic mechanisms. Cell type localization and SSc-specific changes were confirmed by spatial molecular imaging. By offering a comprehensive view of transcriptional dysregulation at single-cell resolution in human esophageal epithelial cells in SSc compared to GERD and healthy tissue, this work clarifies the state of epithelial cells in SSc-induced esophageal dysfunction.

## INTRODUCTION

Systemic sclerosis (SSc), also known as scleroderma, is an immune-mediated rheumatic disease of unknown etiology that causes fibrosis of the skin and internal organs. Although uncommon (18-26 per 100,000 people)^1–4^, SSc is one of the deadliest autoimmune disorders^5^, with a 10-year survival rate of 45-68% following diagnosis^4,6,7^. Over 90% of individuals with SSc report gastrointestinal dysfunction^8^, with esophageal dysmotility being the most common gastrointestinal manifestation^9^. Individuals with SSc and esophageal involvement typically suffer from chronic acid reflux and dysphagia due to loss of esophageal motility and are at much greater risk of esophageal stricture and Barrett’s esophagus^8,10,11^. The prevalence of esophageal involvement was nearly twice as high in individuals with SSc who died within 5 years of disease diagnosis^12^.

There are multiple, interrelated mechanisms that may cause esophageal dysmotility in SSc. Vascular damage and/or neurogenic impairment can lead to smooth muscle atrophy^13^, which is the predominant esophageal pathology in autopsies of patients with SSc^14^. Inflammatory signatures^15^, absence of anti-centromere antibodies^16^, and presence of anti-topoisomerase I antibodies are associated with esophageal pathologies, as well^8^. Complicating research efforts is significant clinical and molecular heterogeneity observed between SSc patients^17–19^ and difficulty distinguishing between effects from autoimmune processes and secondary factors such as severe gastroesophageal reflux disease (GERD). Consequently, the pathogenesis of esophageal disease in SSc remains poorly understood.

Functional investigations of SSc esophageal involvement have primarily focused on peristalsis or contractions of the muscularis propria^20^. However, the mucosa is no less essential for normal esophageal transport^21,22^, and recent studies have suggested that esophageal epithelial cells (EECs) may play a more central role in the pathogenesis of SSc esophageal involvement than previously thought^23^. Mouse models with epithelial-cell-specific knockout of *Fli1*, an Ets transcription factor implicated in SSc pathogenesis^24^, spontaneously develop dermal and esophageal fibrosis and interstitial lung disease^25^. Importantly, the resultant lung disease is mediated by T-cell autoimmunity, but fibrosis of the skin and esophagus persist in mice additionally lacking mature T and B cells, suggesting that the fibrosis is principally driven by the epithelia^23,25^.

Most studies implicating epithelial cells in SSc esophageal dysmotility have been performed in mice or *in vitro*, and molecular analyses in humans have so far been limited to bulk tissues^15^. Furthermore, single-cell gene expression studies performed in other diseases of the esophagus, including squamous cell carcinoma^26,27^ and allergic eosinophilic esophagitis^28,29^, have identified substantial cellular and molecular changes in EECs compared to healthy tissue, such as significant expansion of non-proliferative suprabasal cells^28,29^. Therefore, to comprehensively investigate the effects of SSc on the esophageal epithelium, we performed single-cell RNA sequencing (scRNA-seq) of esophageal mucosal biopsies from SSc patients with clinically significant esophageal involvement and compared EEC distributions and gene expression signatures to non-SSc individuals with GERD and healthy controls (HCs). We further validated cell type localization and SSc effects using spatial molecular imaging from independent SSc and HC samples. By examining gene expression profiles from human tissue at single-cell resolution, this work sheds essential light on the cellular roots of esophageal dysfunction in SSc by clarifying the pathogenic role of the squamous epithelium, one of the most integral tissues supporting healthy esophageal function.

## METHODS

### Subjects

The study protocol received approval from the Northwestern University Institutional Review Board. The SSc study population were recruited from adult patients aged 18 to 89 diagnosed with SSc according to the 2013 ACR/EULAR Classification Criteria, who visited the Northwestern Medicine Esophageal Center between 2020 and 2021for esophageal symptom evaluation and motility testing, as described previously^20^. Patients with non-SSc GERD were recruited at the first clinic visit following positive wireless pH testing^29^. Patients with technically inadequate panometry or manometry studies, previous foregut surgeries (including prior pneumatic dilation), candida esophagitis, history of esophageal cancer, or mechanical obstructions in the esophagus such as esophageal stricture, eosinophilic esophagitis, severe reflux esophagitis (Los Angeles classification C or D), or hiatal hernias larger than 5 cm were excluded. HCs met asymptomatic criteria including lack of esophageal symptoms, no history of alcohol dependency or tobacco use, body mass index less than 30kg/m^2^, and no current antacid or proton pump inhibitor treatment^29^. The study protocol received approval from the Northwestern University Institutional Review Board.

### Symptom evaluation and motility testing

Esophageal function testing included functional lumen imaging probe (FLIP) panometry during sedated endoscopy, high-resolution manometry (HRM), and in some subjects, 24-hour pH-impedance or timed-barium esophagogram, which were completed and interpreted as previously described^30–33^. HRM classifications were based on the application of the Chicago Classification v4.0 to 10 supine and 5 upright test swallows^33^. FLIP panometry classifications were based on previous evaluation of asymptomatic volunteers and patients^34–36^. During endoscopy (same encounter as FLIP), esophageal mucosal biopsies were obtained at approximately 5-cm (“distal esophagus”) and 15-cm (“proximal esophagus”) proximal to the squamocolumnar junction using Boston Scientific Radial Jaw 4 standard capacity forceps. Acid exposure was measured as the percentage of time with esophageal pH<4 over a monitoring period of 24 hours. Most patients also completed validated symptom-questionnaires (patient-reported outcome measures) on the day of esophageal motility testing with FLIP or HRM, including the Brief Esophageal Dysphagia Questionnaire (BEDQ), the Gastroesophageal Reflux Disease Questionnaire (GerdQ), and the Northwestern esophageal quality of life (NEQOL) survey.^37–39^. SSc disease duration was measured from onset of the first non-Raynaud symptom.

### Single-cell RNA-sequencing sample processing and sequencing

Esophageal mucosal biopsies from proximal and distal esophagus were processed for scRNA-seq as described in Clevenger et al^29^. Briefly, following tissue digestion and filtering, cell suspensions met 85% minimum viability via Cellometer Auto2000 (Nexcelom Bioscience). Cells were loaded into a Chromium iX controller (10X Genomics) on a Chromium Next GEM Chip G (10X Genomics) to capture approximately 10,000 cells per sample and were processed for encapsulation following the manufacturer’s protocol. Cell barcoding and library construction were performed using the 10X Genomics Chromium Next GEM Single Cell 3’ Reagents Kits v3.1 and Dual Index Kit TT Set A according to the manufacturer’s protocol. Sequencing was performed by the NUSeq Core using the Illumina NovaSeq 6000. Paired-end reads consisting of a 28-base-pair read for cell barcodes and unique molecular identifiers and a 90-base-pair read for transcripts were aligned to the GRCh38 reference transcriptome using Cell Ranger v6.1.2.

### Single-cell sample and cell quality control

Samples with high ambient RNA (<70% of sequencing reads mapped to cells) were removed from consideration. Gene expression counts generated by Cell Ranger were analyzed using Seurat v4.3.0^40^ in R v4.3.0. Cells with low gene diversity (<200 unique genes), low unique molecular identifier counts (<500), or excessive counts (>100,000) were removed from consideration. Cells with high proportions of mitochondrial DNA reads, indicative of apoptosis or lysis, were removed using the bivariate regression models implemented in the miQC package^41^. Cells were filtered if their proportion of reads mapped to mitochondrial DNA exceeded 0.05 and their posterior probability of belonging to a compromised cell distribution was greater than 0.75 according to miQC. Doublets, in which more than one cell is clumped together in a single droplet, were predicted and removed using scDblFinder^42^ with an expected, additive doublet rate of 0.01 per thousand cells with standard deviation of 0.01.

### Sample integration and cell type annotation

Individual scRNA-seq expression profiles were normalized and integrated on 3,000 variable features using Seurat’s SCTransform procedure^43^ with v2 regularization^44^ and canonical correlation analysis dimension reduction. Integration anchors were determined using a reference consisting of three pairs of proximal and distal samples from HCs. Principal component (PC) analysis was performed on the transformed and integrated expression counts. Uniform manifold approximation and projection (UMAP)^45^ embeddings were calculated on the top 40 PCs with n=200 neighbors. Cell clustering was performed using Seurat’s modularity-based clustering on a shared nearest neighbor (SNN) graph with k=25 on the top 40 PCs. Clusters were annotated according to the expression of established cell-specific gene markers (**Supplemental Figure 1**). For annotation of smaller cell clusters, we first filtered out the epithelial, myeloid, lymphoid, and endothelial cell clusters and then recomputed PCs and re-clustered the subset (PCs = 25, k=15).

### Epithelial cell characterization

After isolating the subset of epithelial cell clusters, we further filtered cells with disproportionately low read counts (≤3,500; **Supplemental Figure 2, A-E**). The samples were then re-integrated using the procedure described above. UMAP embeddings were calculated on the top 35 PCs with n=100 neighbors. Epithelial compartments (basal, proliferating basal, proliferating suprabasal, suprabasal, and superficial) were identified according to expression of established compartment-specific and cell cycle markers (**Supplemental Figure 2F**)^28,29,46–48^. For the basal, suprabasal, and superficial compartments, we computed composite gene signature scores using Seurat’s AddModuleScore() function (**Supplemental Figure 2G**). Cells were initially clustered on an SNN graph with k=50 with resolution=0.25 (**Supplemental Figure 2H**), but because the modularity optimization clustering implemented in Seurat grouped proliferating cells across different epithelial compartments and inconsistently distinguished suprabasal and superficial cells, we applied K-means clustering on the expression of the compartment-specific markers (**Supplemental Figure 2F**). We performed separate K=2 K-means clustering on 1) the Seurat clusters containing basal and early suprabasal cells (clusters 1-5) to distinguish basal from suprabasal compartments and 2) the Seurat clusters containing late suprabasal and superficial cells (clusters 6-8) to demarcate suprabasal and superficial compartments (**Supplemental Figure 2, I-J**). Proliferating epithelial cells were identified using Seurat’s CellCycleScoring() function. Cells predicted as being in S or G2/M phases based on the relative expression of corresponding markers^49^ were labeled as proliferating cells (**Supplemental Figure 2K**). We tested for statistical differences in proportions of epithelial cell populations between sample groups using MASC^50^.

### Gene expression analysis

To identify dysregulated pathways in SSc, we first calculated differential gene expression between conditions (SSc, non-SSc GERD, HCs) within each epithelial compartment, considering each cell as a sample. Differential gene expression was calculated using the FindMarkers() function in Seurat with the Wilcoxon rank-sum test on log-normalized RNA counts for genes expressed in at least 1% of cells by compartment. We adjusted for multiple testing using a Bonferroni correction accounting for the number of genes tested. Genes with an adjusted P<0.05 and absolute log_2_ fold change >0.1 were considered significantly differentially expressed. ^51,52^ Genes were then ranked by the magnitude and statistical certainty of their expression differences:

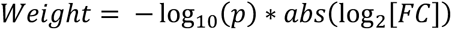

Gene set enrichment was calculated using 3,795 canonical pathway gene sets from the Human Molecular Signatures Database^53^ using gene set enrichment analysis (GSEA)^51,52^. Statistical gene set enrichment was calculated using one-way positive enrichment with 100,000 permutations on pathways with at least 10 genes. Mitochondrial and ribosomal genes were excluded from the GSEA.

For pseudobulk differential expression analysis we aggregated RNA counts per sample within epithelial compartments. Genes with at least one count per million detected in at least 75% of samples in at least one condition per epithelial compartment were analyzed. To mitigate outlier-driven signals, genes with log-transformed expression greater than 3 standard deviations from the mean in exactly one individual were excluded. Differential gene expression was evaluated using edgeR v3.42.4^54–56^. Gene expression was modeled using a quasi-likelihood negative binomial generalized linear model, with global negative-binomial trended dispersion estimated across all samples and gene-specific quasi-likelihood dispersions estimated within each epithelial compartment. Differences in gene expression across conditions were assessed using quasi-likelihood F-tests with false-discovery rate (FDR) adjustment. Differences in the proportions of genes that were significantly differentially expressed between groups were assessed using a chi-square test. Differences in gene expression profiles between biopsy locations were determined by measuring intra-individual correlations for the 2,000 most variable genes at the single-cell level aggregated by sample, epithelial compartment, and biopsy location.

Transcription factor enrichment analyses were conducted on the pseudobulk results using the enricher() function from clusterProfiler v4.8.3^57^. We used consensus results from ENCODE^58^ and ChEA^59^, as curated by the Ma’ayan Lab^60^ and available at http://amp.pharm.mssm.edu/Enrichr/ ^61^ to test for the enrichment of transcription factor target genes from among genes that were differentially expressed between SSc, GERD, and HCs with Bonferroni-adjusted p<0.05. Background gene sets were comprised of the genes included in the pseudobulk differential expression analyses for each epithelial compartment. Transcription factors absent from the background gene set were removed as candidate transcription factors. Separate enrichment tests were performed for all differentially expressed genes and those that were upregulated or downregulated in SSc. Enrichment results were adjusted using a Bonferroni correction accounting for the number of transcription factors with more than 2 genes that were significantly differentially expressed.

### Pseudobulk sample permutation

Because of the sample size imbalance by disease state, which inherently yields different statistical power for different tests across conditions, we permuted differential expression testing with equivalent sample sizes to better evaluate the relative extent of dysregulation by EEC compartment and biopsy location between SSc and GERD. We permuted all possible combinations of SSc samples with sample size n=4 for both the proximal and distal regions and used the median statistics in the SSc vs. HC tests to more directly compare the results with those observed in GERD vs. HCs.

### Spatial imaging sample preparation

Tissue sections were mounted on Leica Biosystems Apex BOND Superior Adhesive Slides (cat# 3800040) and subsequently baked overnight at 60 °C, followed by preparation for in-situ hybridization (ISH) by deparaffinization in CitriSolv (2x 5min), 100% ethanol (2x 5min) and heat-induced epitope retrieval (HIER) at 100 °C for 15 min using low pH citrate buffer (NanoString/Bruker). After HIER, tissue sections were digested with Proteinase K (2 µg/ml) diluted in PBS at 40 °C for 30 min. Tissue sections were then washed twice with diethyl pyrocarbonate (DEPC)-treated water (DEPC H2O) and incubated in 0.0015% fiducials (NanoString/Bruker) in 2X saline sodium citrate, 0.001% Tween-20 (SSCT solution) for 5 min at room temperature in the dark. Excess fiducials were rinsed from the slides with 1X PBS, then tissue sections were fixed with 10% neutral buffered formalin (NBF) for 5 min at room temperature. Fixed samples were rinsed twice with Tris-glycine buffer (0.1 M glycine, 0.1 M Tris-base in DEPC H2O) and once with 1X PBS for 5 min each before blocking with 100 mM N-succinimidyl (acetylthio)acetate (NHS-acetate, Thermo Fisher Scientific) in NHS-acetate buffer (0.1 M NaP, 0.1% Tween pH 8 in DEPC H2O) for 15 min at room temperature. The sections were then rinsed with 2X saline sodium citrate (SSC) for 5 min and an Adhesive Hybridization Chamber (NanoString/Bruker) was placed over the tissue. ISH probes were prepared by incubation at 95 °C for 2 min and submerged in ice, and the ISH probe mix was pipetted into the hybridization chamber. The chamber was sealed to prevent evaporation, and hybridization was performed at 37 °C for 18 hrs. Tissue sections were washed twice in 50% formamide (Ambion) in 2X SSC at 37 °C for 25 min, washed twice with 2X SSC for 2 min at room temperature, and blocked with 100 mM NHS-acetate in the dark for 15 min. In preparation for loading onto the spatial imaging instrument, a flow cell was affixed to the slide.

### Spatial molecular imaging and data processing

Single-cell spatial transcriptomic analysis was performed using the CosMx Spatial Molecular Imager (NanoString/Bruker), featuring the 6k Human Discovery Panel gene list, which consists of 6,175 RNA targets designed to provide coverage across all major biological pathways. Spatial fields of view (FOVs) with average counts per cell <100 were removed from consideration. Cells with low RNA counts (<20), high proportions of counts from negative probes (>0.1), extreme size (p<0.01 by Grubb’s test), or more detected genes than counts were removed. Gene probes with expression levels below median background noise, as determined by negative probe detection, were also removed. Counts were log normalized using Seurat^40^. Cell segmentation errors were predicted using scDblFinder^42^. To annotate the cells, we performed label transfer from the single-cell dataset with SingleR^62^ using default parameters on the intersect of quantified genes. Gene expression differences between groups in the spatial data were evaluated using Wilcoxon rank-sum tests.

### Clinical phenotype associations

To test for associations with categorical measures of esophageal function (FLIP, HRM, and a combined FLIP and HRM with neuromyogenic [NM] classification^63^), we performed ordinal logistic regression that modeled likelihoods of increasing clinical severity. For this analysis, we excluded SSc patients with achalasia or esophagogastric junction outflow obstruction (EGJOO). For FLIP we combined the “borderline/diminished” and “impaired/disordered” phenotypes. For NM we combined the Stage I and Stage II ineffective motility phenotypes. The resultant ordering for the ordinal regression was “normal” < “ineffective” < “absent” for HRM; “normal” < “borderline/diminished & impaired/disordered” < “absent” for FLIP; and “normal” < “stages I & II – ineffective” < “stage III – absent” for NM.

For association testing with quantitative clinical measures in SSc, we first performed a principal components analysis (PCA) on a set of nine quantitative clinical traits: HRM mean distal contractile interval (DCI), HRM basal esophagogastric junction (EGJ) pressure, HRM EGJ contractile index, HRM median integrated relaxation pressure (IRP), FLIP intra-balloon pressure at 60ml, FLIP EGJ distensibility index, the GerdQ impact score, the BEDQ score, and the NEQOL score. Missing values were imputed with the mean across all patients. We then modeled associations with the first two PCs using linear regression. Gene expression was represented using log-transformed counts.

## RESULTS

Whole tissue scRNA-seq was performed on paired proximal and distal mucosal biopsies from ten patients with SSc, four non-SSc patients with GERD, and six HCs. All participants but one in each group were female (**Table 1**). SSc patients were significantly older than HCs (54.9 vs 27.0 years; P<0.001). Among SSc patients, six had the limited cutaneous subtype, three had the diffuse cutaneous subtype, and one had sine scleroderma with no cutaneous symptoms (**Supplemental Table 1**). Mean SSc disease duration at the time of biopsy was 132 months. All SSc patients had positive serum antinuclear antibodies and impaired esophageal motility. Following sample-level quality control (**Supplemental Table 2**), 39 samples were retained for analysis, including 19 from the proximal esophagus and 20 from the distal esophagus. Four additional sets of proximal and distal biopsies were collected for spatial molecular imaging from two SSc patients with diminished esophageal contractility and two healthy controls (**Supplemental Table 3**).

**Table 1.** Sc-RNA-seq patient demographics and clinical characteristics.

|  | HCs |  | SSc |  | GERD |  |
| --- | --- | --- | --- | --- | --- | --- |
| Demographics | N = 6 |  | N = 10 |  | N = 4 |  |
| Sex – female | 5 (83.3%) |  | 9 (90%) |  | 3 (75%) |  |
| Race – white | 5 (83.3%) |  | 8 (80%) |  | 2 (50%) |  |
| Ethnicity – non-Hispanic | 5 (83.3%) |  | 8 (80%) |  | 2 (50%) |  |
| Race/ethnicity – unknown/NR | 1 (16.7%) |  | 2 (20%) |  | 2 (50%) |  |
| Traits | N | Median (IQR) | N | Median (IQR) | N | Median (IQR) |
| Age (yrs) | 6 | 27 (25-29) | 10 | 54 (47-70) | 4 | 42 (30-56) |
| BMI (kg/m<sup>2</sup>) | 6 | 23 (22-25) | 10 | 22 (19-30) | 4 | 24 (23-36) |
| **Esophageal motility** |  |  |  |  |  |  |
| Mean DCI (mmHg•s•cm) | 6 | 464.2 (386-6-1026.1) | 10 | 57.5 (0.0-206.1) | 2 | 509.6 (367.4-651.7) |
| Pressure 60ml (mmHg) | 6 | 43.5 (38.3-54.5) | 9 | 31.0 (30.5-40.3) | 0 | - |
| **EGJ barrier function** |  |  |  |  |  |  |
| End-expiratory EGJ pressure (mmHg) | 6 | 7 (4-8) | 10 | 6 (3-10) | 2 | 12 (11-12) |
| EGJ contractile index (mmHg•cm) | 6 | 19.4 (15.2-29.0) | 10 | 21.0 (19.1-27.5) | 2 | 19.0 (10.8-27.1) |
| Median IRP (mmHg) | 6 | 4 (3-9) | 10 | 6 (4-12) | 2 | 8 (6-9) |
| EGJ-DI (mm<sup>2</sup>/mmHg) | 6 | 6.2 (5.2-8.5) | 9 | 6.3 (4.1-9.2) | - | - |
| **Reflux severity** |  |  |  |  |  |  |
| AET (%) | 6 | 0.8 (0.5-2.8) | 4 | 8.7 (3.3-14.5) | 3 | 16.8 (13.9-17.3) |
| GerdQ<sup>118</sup> score | 6 | 0 (0-0) | 10 | 3 (1-4) | 3 | 3 (3-4) |
| BEDQ<sup>38</sup> score | 6 | 0 (0-0) | 8 | 8 (3-14) | 3 | 1 (1-1) |
| NEQOL<sup>39</sup> score | 6 | 56 (56-56) | 8 | 31 (23-47) | 3 | 36 (29-46) |

AET, acid exposure time; BEDQ, brief esophageal dysphagia questionnaire; BMI, body mass index; DCI, distal contractile integral; DI, distensibility index; EGJ, esophagogastric junction; HCs, healthy controls; IQR, inter-quartile range; IRP, integrated relaxation pressure; NEQOL, Northwestern esophageal quality of life; NR, not reported; scRNA-seq, single-cell RNA sequencing.

### Characterization of esophageal mucosal cell populations

A total of 306,372 esophageal cells (7,856 mean cells per sample) were retained for cell clustering and gene expression analysis. Each cell type was identified according to the expression of established cell-specific gene markers including *KRT6A, KRT13,* and *KRT15* for epithelial cells (**Figure 1, A-D; Supplemental Table 4**; **Supplemental Figure 1**). Epithelial cells comprised the bulk of the sample (n=264,858; 86.45%), followed by lymphoid cells (n=25,129; 8.20%), myeloid cells (n=10,019; 3.27%), endothelial cells (n=4,027; 1.31%), and all other cell types (n=2,339; 0.76%). We observed heterogeneity in non-epithelial cell type proportions by condition and biopsy location (**Figure 1E**) but focused on the epithelium for the remainder of the study.

**Figure 1.**
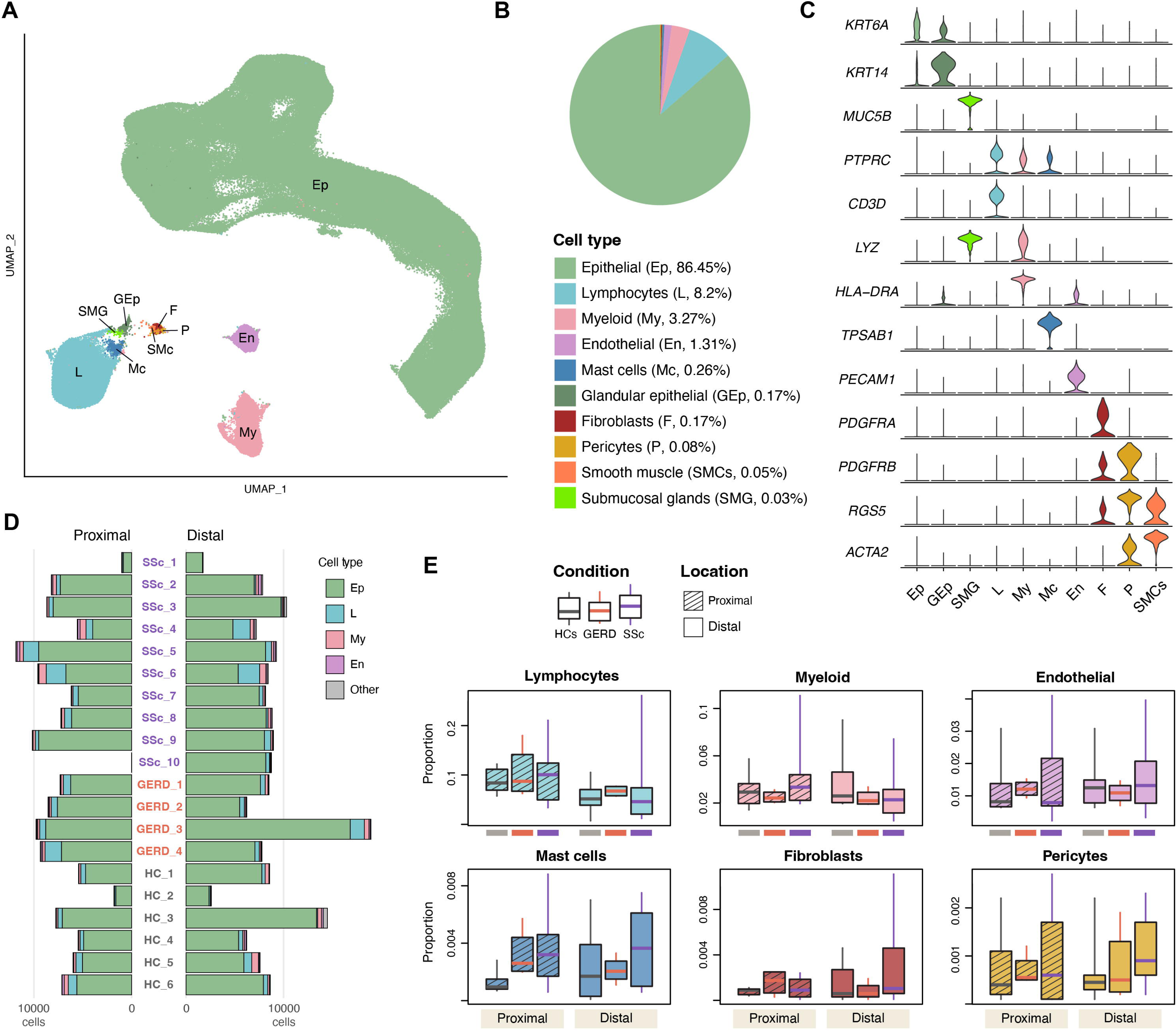
ScRNA-seq sample composition by cell type. **A)** Integrated UMAP embedding of all cells (n=306,372) labeled by cell type. **B)** Proportion of cells by cell type. **C)** Expression by cell type of canonical markers. **D)** Sample-wise (n=39) distribution of major cell types. **E)** Proportion of major non-epithelial cell types by condition and biopsy location. Box whiskers extend to range of values.

### Significant loss of superficial EECs in SSc

Following isolation of the epithelial cell cluster and additional quality control (**Supplemental Figure 2, A-E**), we characterized the remaining 230,720 EECs according to their differentiation and cell cycling states (**Figure 2A**). We classified the EECs into five primary compartments: basal (n=55,818; 24%), proliferating basal (n=36,439; 16%), proliferating suprabasal (n=25,314; 11%), suprabasal (n=82,283; 36%), and superficial (n=30,866; 13%) based on relative expression of canonical epithelial genes and cell cycle markers (**Figure 2, B-C; Supplemental Figure 2, F-K**). Spatial localization of the annotated single-cell clusters was confirmed using spatial molecular imaging (**Figure 2D, Supplemental Figure 3**). Roughly 40% of basal cells and 24% of suprabasal cells were proliferating, and the fractions of cells that were proliferating did not differ by disease (**Figure 2E**). Proportions of basal and suprabasal cells were not significantly different across conditions, but there were significantly fewer proximal superficial cells in SSc compared to HCs (µ_SSc_=0.10, µ_HC_=0.21, P=0.003, **Figure 2E**).

**Figure 2.**
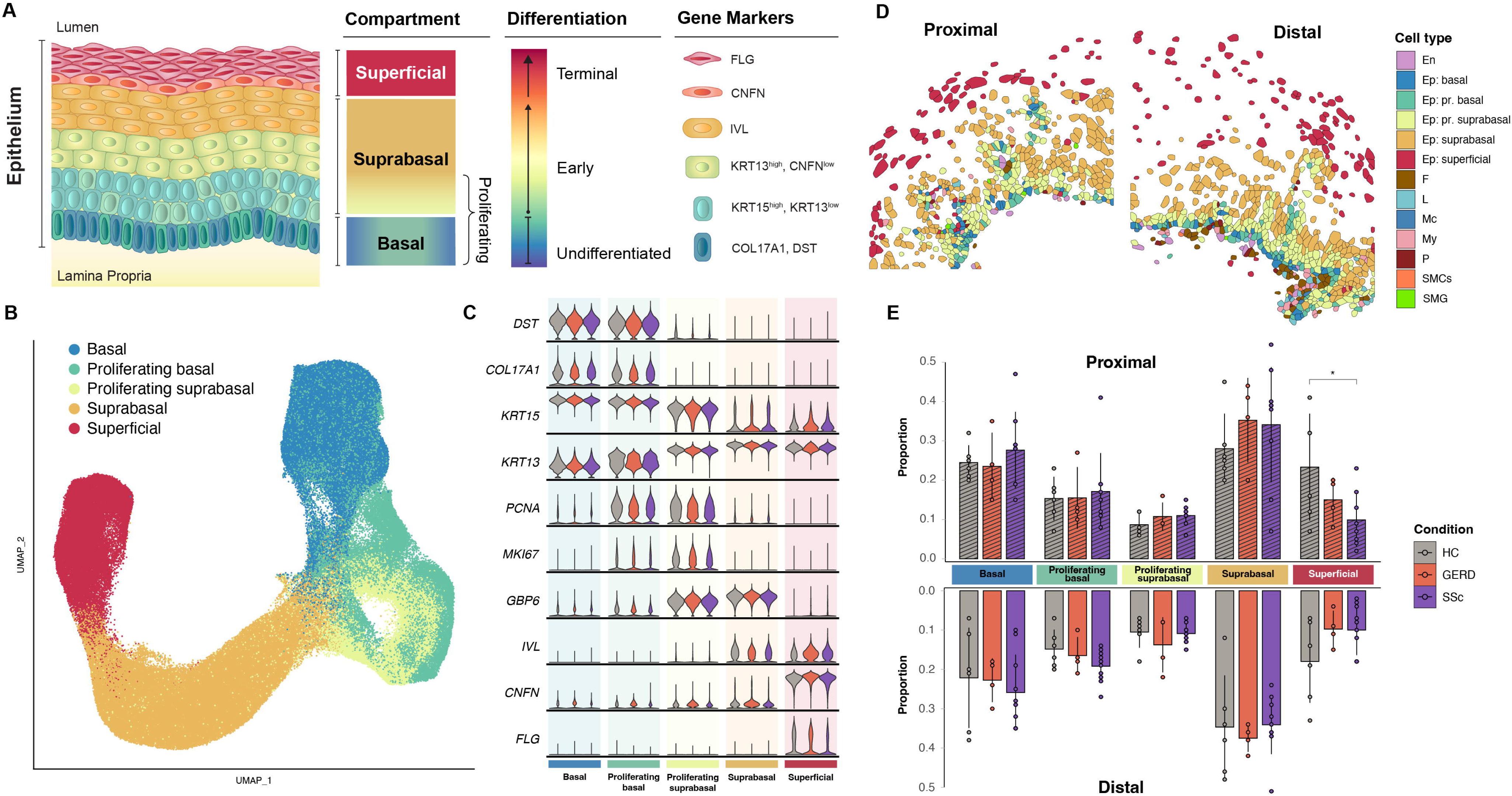
Landscape of esophageal epithelial cells. **A)** Histological summary of the human esophageal epithelium, adopted from Clevenger et al.^29^ Alternatively shaded cells within the same layer represent proliferating cells. NOTE: The layers of replicating cells not attached to the basement membrane, labeled here as proliferating suprabasal cells, are sometimes referred to as epibasal cells^29^. **B)** Integrated UMAP embedding of all esophageal epithelial cells (EECs, n=230,720) labeled by epithelial compartment. **C)** Expression of EEC compartment and cell cycle markers by condition. **D)** CosMx spatial molecular imaging of proximal and distal esophageal mucosal tissue in scleroderma with absent contractility, annotated using label transfer from the scRNA-seq data. **E)** Proportions of cells by condition, EEC compartment, and biopsy location. The bars denote the mean values, the vertical lines the standard deviations, and the points the individual sample proportions. Pairwise differences across conditions were evaluated statistically using MASC^50^. *p<0.05.

### EEC gene dysregulation predominantly in superficial cells, highly correlated in SSc and GERD

Most differentially expressed genes (DEGs) were only differentially expressed in one epithelial compartment (**Supplemental Figure 4A**), and differential gene expression between conditions was most pronounced in superficial EECs (**Figure 3A**). At the single-cell level, 674 and 434 genes were significantly differentially expressed between SSc and HCs in the superficial compartment in the proximal and distal regions, respectively, compared to 171 proximal and 157 distal non-proliferating basal DEGs and 172 proximal and 377 distal non-proliferating suprabasal DEGs (**Supplemental Tables 5-6**). To more robustly characterize gene expression differences between conditions^64^, we performed pseudobulk differential expression analysis by aggregating single-cell RNA counts per sample within EEC compartments. Overall, pseudobulk gene expression patterns clustered by compartment rather than by condition or biopsy location (**Supplemental Figure 4B**). Significant differences in gene expression between conditions were largely limited to superficial cells, suggesting that the apical cell layers of the esophageal epithelium are most affected by SSc (**Supplemental Figure 4, C-D**). There were 3,572 genes differentially expressed between SSc and HCs with FDR q<0.05 in the superficial compartment, compared to just 232 total in all other compartments (**Supplemental Table 7**).

**Figure 3.**
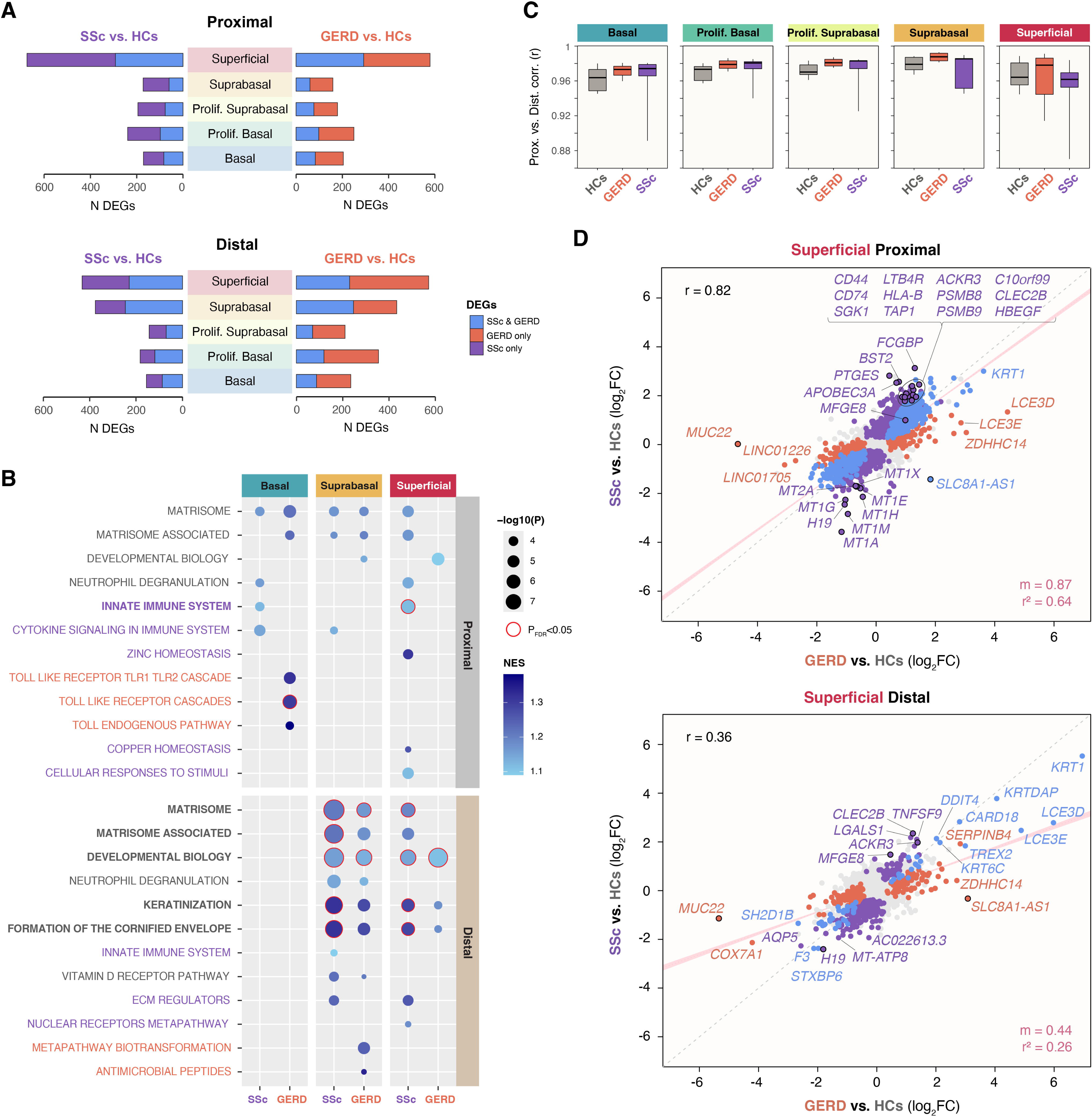
Differential gene expression between conditions in esophageal epithelial cells. **A)** The number of significantly differentially expressed genes at the single-cell level with |log_2_FC| >0.1 are shown for SSC vs. HCs (purple) and GERD vs. HCs (orange), by epithelial compartment and biopsy location. Blue bars account for genes that were differentially expressed in both SSc and GERD compared to HCs. **B)** Gene set enrichment analysis results, showing all pathways enriched with P<0.001 (unadjusted) for SSc vs. HCs (purple) and GERD vs. HCs (orange), split by non-proliferating epithelial compartment and biopsy location. Points circled in red indicate statistical significance after adjusting for the number of tested pathways (P_FDR_<0.05). Pathways in bold denote pathways with significant enrichment in SSc. **C)** The distribution of intra-sample EEC gene expression correlations for the 2,000 most variable genes between proximal and distal biopsies are shown by epithelial compartment and condition. Box whiskers extend to range of values. **D)** Pseudobulk gene expression differences in log_2_FC are shown for SSc vs. HCs (Y axis) against GERD vs. HCs (X axis) within the superficial compartment in the proximal and distal esophagus regions. Significant DEGs in both conditions are highlighted in blue, significant DEGs in SSC only are highlighted in purple, and significant DEGs in GERD only are highlighted in orange. The correlation between comparisons is shown in the upper left, and the slope and coefficient of determination for the modeled linear regression with intercept=0 is displayed in the lower right. Trendlines with 95% confidence intervals are plotted in pink, and the dashed grey lines denote where y=x. Encircled points highlight disease-specific DEGs that are referenced in the main text and plotted in **Supplemental** Figure 4H, which shows the distribution of expression aggregated by sample in the superficial compartment across conditions and biopsy locations.

The differential expression observed in SSc was strongly correlated with the changes seen in GERD, with many of the same genes significantly dysregulated in both SSc and GERD compared to HCs. Overall correlations in pseudobulk log_2_ fold change between SSc vs. HCs and GERD vs. HCs were consistently strong, ranging between r=0.51-0.62 across EEC compartments (**Supplemental Table 7**). Depending on the compartment and biopsy location, 22-44% of single-cell DEGs in SSc or GERD were differentially expressed in both conditions (**Figure 3A**; **Supplemental Table 5**). Consequently, the single-cell DEGs in SSc and GERD were significantly enriched for some of the same pathways, particularly those related to the extracellular matrix and keratinization (**Figure 3B**). These included matrisome (suprabasal, distal; superficial, distal) and matrisome-associated genes (suprabasal, distal); cornified envelope formation (suprabasal, distal; superficial, distal), and keratinization genes (suprabasal, distal; superficial, distal); developmental biology-related genes (suprabasal, distal; superficial, distal). The leading edges of these significant pathways, which comprised the genes driving their enrichment, included shared protein families, including serine protease inhibitors (serpins), keratins, small proline-rich proteins (SPRRs) and S100 proteins (**Supplemental Table 8**). Among the most enriched pathways specific to SSc were those related to the regulation of metal and immune homeostasis. In particular, innate immune system genes were uniquely enriched in SSc compared to HCs in superficial EECs of the proximal esophagus.

Overall gene expression levels were highly similar between esophageal biopsy locations, with mean intra-individual correlations grouped by condition and epithelial compartment ranging from r=0.95 to r=0.99 for the 2,000 most variable genes (**Figure 3C**; **Supplemental Table 9**). However, we did observe some notable differences in differential gene expression patterns by biopsy location. For example, single-cell differential expression in the suprabasal compartment was significantly greater in the distal esophagus than in the proximal esophagus for both SSc (172 proximal vs. 377 distal, p=2.5×10^-20^) and GERD (159 proximal vs. 434 distal, p=1.1×10^-32^; **Figure 3A; Supplemental Figure 4A**). Furthermore, at the single-cell level, there were more unique DEGs in SSc vs. HCs in the proximal esophagus (897 proximal vs. 806 distal, p=7.0×10^-2^), but the opposite was true in GERD vs. HCs (910 proximal vs. 1,075 distal, p=1.6×10^-5^; **Figure 3A**). Similarly, within the superficial compartment, where most differential expression was observed, GERD exhibited greater relative changes in pseudobulk gene expression in the distal esophagus than in the proximal region, as indicated by the decreased slope between fold-changes (m=0.44 vs. 0.87; **Figure 3D; Supplemental Table 10**). The superficial compartment also featured the greatest range and lowest overall values of intra-individual gene expression correlations between biopsy locations, particularly in SSc (**Figure 3C**). These differences by biopsy location suggest that the relative disease effect on the proximal esophagus compared to the distal esophagus is greater within SSc than within GERD.

More genes were significantly differentially expressed in SSc than in GERD in the pseudobulk analysis, but in terms of relative fold change vs. HCs, we observed greater expression changes in GERD (**Figure 3D**). To determine whether the greater number of SSc samples were driving this discrepancy, we performed all possible sample permutations with equal numbers of SSc and GERD samples (n=4) and repeated differential gene expression testing to more directly compare the relative number of DEGs in both conditions. In superficial cells from the proximal esophagus, there were more DEGs in SSc in 60% of permutations, but the relative FC differences were greater in GERD in 61% of permutations (**Supplemental Figure 4, E-F; Supplemental Table 11**). In the distal region, there were more DEGs in GERD in 62% of permutations, and the relative FC differences were greater in GERD in all permutations. The correlations between SSc and GERD in terms of relative gene expression changes were consistently much higher in the proximal region permutations than in the distal region (**Supplemental Figure 4G**). The permutation results confirmed that the differences in gene expression observed in SSc relative to GERD were not simply due to differences in sample size. In summary, the superficial epithelium is most altered in SSc, exhibiting similar transcriptional changes to GERD, but with relatively greater effect in the proximal esophagus compared with distal.

### SSc-specific gene dysregulation points to immunoregulatory pathways

Among the most upregulated SSc-specific genes (**Figure 3D; Supplemental Figure 4H**) were genes related to inflammation (*PTGES*, *MFGE8*)^65,66^, innate immune response (*FCGBP, BST2*, *CD44*, *APOBEC3A*)^67–70^, immune cell migration (*C10orf99*, *LTB4R*, *ACKR3*)^71–73^, antigen presentation (*HLA-B*, *CD74*, *TAP1*, *PSMB8*, *PSMB9*)^74^, natural killer cell activation (*CLEC2B*)^75^, and fibroproliferation (*SGK1*, *HBEGF*)^76,77^. Four of the top five and eight of the top 20 most downregulated SSc-specific genes by fold change in superficial EECs in the proximal esophagus were metallothioneins (*MT1A*, *MT1E*, *MT1F*, *MT1G*, *MT1H*, *MT1M*, *MT1X*, *MT2A*). The metallothioneins comprised the bulk of three pathways uniquely enriched in SSc with P<0.001: zinc homeostasis, copper homeostasis, and cellular responses to stimuli (**Figure 3B**). The most down-regulated SSc-specific gene across both the proximal and distal esophagus was the *H19* lncRNA. Interestingly, the most down-regulated gene by fold change in GERD, *MUC22*, was not differentially expressed in SSc. One gene, *SLC8A1-AS1*, was significantly differentially expressed in both GERD and SSc, but in opposite directions (**Figure 3D; Supplemental Figure 4H; Supplemental Table 10**).

### Transcription factors enriched in proximal SSc esophagus

We next examined whether DEGs in SSc superficial cells were significantly enriched for targets of specific transcription factors. Among genes differentially expressed in SSc compared to GERD, the target genes of IRF1 were significantly enriched (**Supplemental Table 12**). IRF1 targets were significantly enriched among all proximal superficial DEGs (p_adj_=0.021) and among only those that were downregulated (p_adj_=0.0024). However, *IRF1* was not itself differentially expressed in superficial cells in SSc compared to GERD. Compared to HCs, three transcription factors were significantly enriched in SSc in the proximal esophagus, including MYC (p_adj_=2.1×10^-7^) and E2F4 (p_adj_=5.5×10^-5^) for upregulated DEGs and NFE2L2 for downregulated DEGs (p_adj_=7.6×10^-4^). *MYC* and *E2F4* were also themselves differentially expressed in SSc compared to HCs in superficial cells (p_adj_=7.8×10^-5^, p_adj_=9.8×10^-4^, respectively). MYC was the most enriched transcription factor in genes upregulated in GERD compared to HCs, but the enrichment was not significant after adjusting for multiple testing (p_adj_=0.11). No transcription factors were significantly enriched in any pairwise comparison in the distal esophagus.

### Fewer metallothioneins and metallothionein-expressing cells in superficial EECs in SSc

With SSc-specific dysregulation most prominent in the superficial cells of the proximal esophagus, we performed sub-clustering of these cells to further examine corresponding expression patterns. We identified five distinct clusters (**Figure 4A**) with unique expression marker patterns (**Figure 4B, Supplemental Figure 5A, Supplemental Table 13**) and varying degrees of differentiation (**Figure 4C**). We then performed label transfer onto the spatially mapped cells to confirm the physical distribution of these clusters within the esophageal epithelium. Clusters 1 and 3 represented the apical, terminally differentiated epithelial layers, as evidenced by their *FLG* expression^78–80^ and spatial localization (**Figure 4D**), but cluster 3 had uniquely high expression of metallothioneins (**Figure 4B, Supplemental Figure 5A**). Cluster 2 was distinguished by its relatively high *CTSV* expression, while clusters 4 and 5 were the least differentiated of the superficial cells and were distinguished by their relative expression of *GJB6* and *FGFBP1*, respectively. Cluster 3 was less abundant in SSc compared to HCs with P<0.05, while no other clusters demonstrated significantly different abundances between SSc and HCs (**Figure 4E**). Metallothionein expression was also significantly lower in SSc compared to HCs or GERD in each cluster, except for cluster 5 (P<5×10^-14^, **Figure 4, F-G**). These observations were reproduced in the spatial imaging data, where cluster 3 was half as abundant in SSc (14.5%) as in HCs (31.6%, **Figure 4D**) and metallothionein expression was likewise significantly lower in SSc compared to HCs in clusters 1-4 (P<1.5×10^-7^, **Figure 4, H-I**). Moreover, metallothionein expression was not significantly different between SSc and HCs in distal superficial cells in the spatial imaging data (**Figure 4I**).

**Figure 4.**
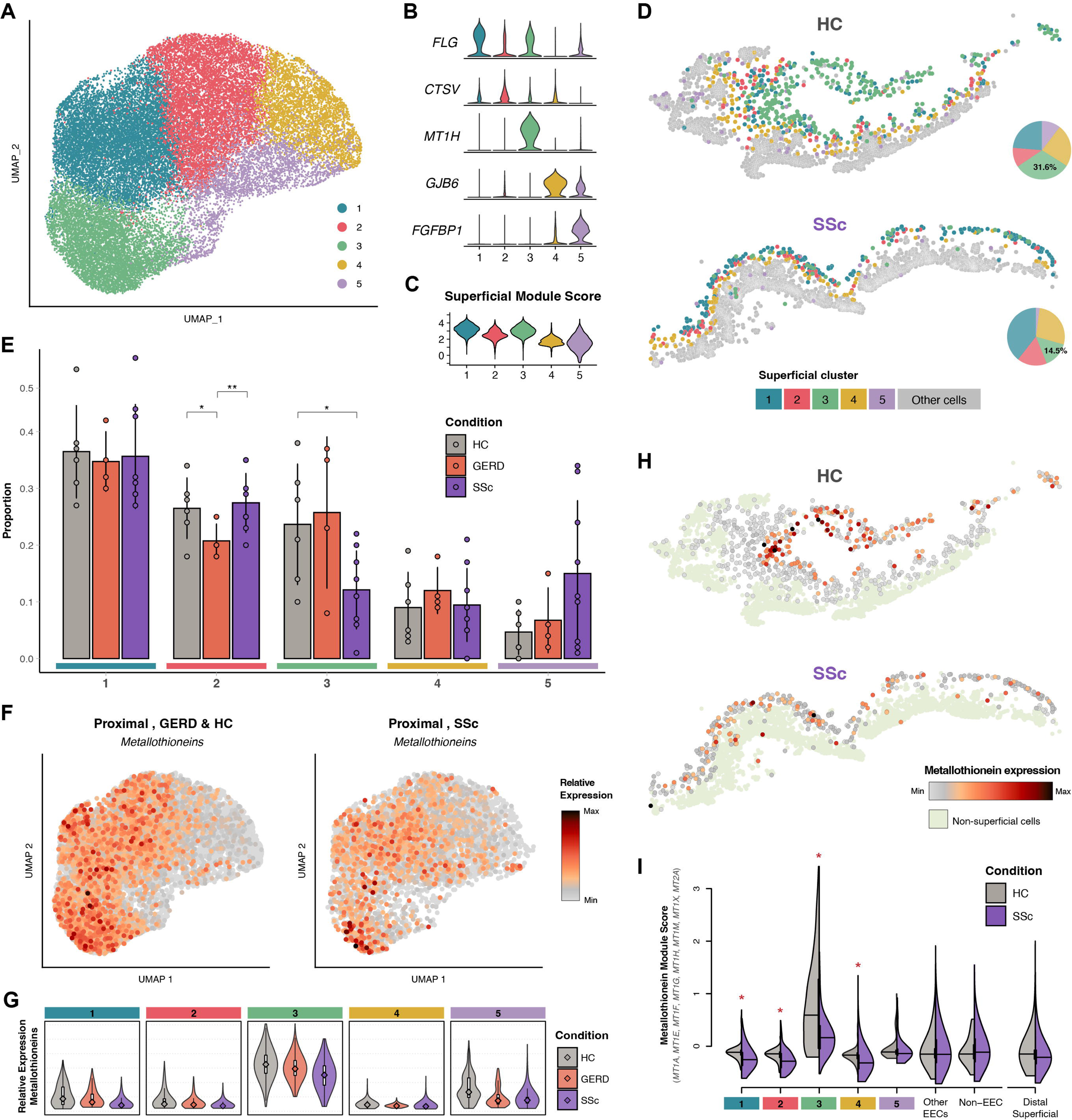
Landscape of superficial esophageal epithelial cells. **A)** Integrated UMAP embedding of all superficial EECs (n=30,866) clustered by unique transcriptional signatures. **B)** Relative expression of cluster-specific gene signatures. **C)** Superficial module score by superficial cell cluster. **D)** CosMx spatial molecular imaging of proximal esophageal mucosal tissue from a healthy control donor and in scleroderma with absent contractility, annotated using label transfer from the scRNA-seq data. Superficial cluster proportions were derived from all imaged proximal tissue (n=2 SSc, 2 HCs). **E)** Proportion of superficial cells by condition and cluster from the proximal esophagus. The bars denote the mean values, the vertical lines the standard deviations, and the points the individual sample proportions. Pairwise differences across conditions were evaluated statistically using MASC^50^. *p<0.05; **p<0.005. **F)** Distribution of metallothionein module score in proximal, superficial cells by condition. The metallothionein module score included *MT1A*, *MT1E*, *MT1F*, *MT1G*, *MT1H*, *MT1M*, *MT1X*, and *MT2A*. **G)** Distribution of relative metallothionein expression by cluster and condition in superficial cells from proximal esophagus. All pairwise comparisons between SSc and HCs were statistically significant, except for cluster 5. **H)** CosMx spatial molecular imaging of proximal esophageal mucosal tissue from a healthy control donor and in scleroderma with absent contractility. Metallothionein module scores are plotted within the superficial EECs. **I)** Distributions of metallothionein module scores by cell cluster and condition from the CosMx spatial molecular imaging data. Clusters withs significant pairwise differences (Wilcoxon) in module score levels (P_FDR_<0.05) are denoted with an asterisk.

Regarding the expression of the significantly enriched transcription factors (IRF1, MYC, E2F4, and NFE2L2; **Supplemental Table 12**) and their target genes, we observed several patterns (**Supplemental Figure 5, B-E**). For *MYC*, *E2F4*, and *NFE2L2*, each gene’s expression was highest on average in cluster 5, the least differentiated superficial cluster (**Figure 4C**), and by condition was highest in SSc, followed by GERD. Aggregate expression of target genes was positively correlated with transcription factor expression for *MYC* (r=0.42) and *E2F4* (r=0.24) and negatively correlated for *IRF1* (r=-0.20) and *NFE2L2* (r=-0.18). Correspondingly, cluster 5 featured the highest target gene expression for *MYC* and *E2F4* and lowest for *NFE2L2*, and target gene expression differences relative to HCs were likewise correlated in SSc and GERD but were more pronounced in SSc. Notably, several of the immune-related genes uniquely upregulated in SSc (**Figure 3D; Supplemental Figure 4H**) displayed a similar expression pattern within superficial cells as *MYC* and *E2F4*, with expression unique to cluster 5 (**Supplemental Figure 5F**, **Supplemental Table 13**). For *IRF1*, however, expression of the transcription factor and its target genes changes differed more in GERD than in SSc relative to HCs.

### *FLI1* not expressed in human EECs

Deletion of the *Fli1* gene in epithelial cells in mice has been shown to recapitulate histological and molecular features of esophageal involvement in SSc^25^. Therefore, we investigated the expression of *FLI1* in our human esophageal samples. The expression of *FLI1* in human EECs was negligible and did not meet the minimum expression thresholds for inclusion in our differential gene expression analysis. We did observe low *FLI1* expression levels in endothelial cells (**Supplemental Figure 6A**), but the expression differences between SSc, HCs, and GERD was not significant (**Supplemental Figure 6B**). There were many FLI1 downstream targets among the proximal, superficial DEGs between SSc and HCs and seven between SSc and GERD (*ARPC2*, *RAB24*, *MTPN*, *PPIF* upregulated; *CEBPZ*, *TRMT10C*, *TBP* downregulated); however, the proportion of these genes among all DEGs was not different than what would be expected by chance (P>0.05), and the average expression of FLI1 targets was higher in GERD and lower in HCs compared to the expression in SSc (**Supplemental Figure 6C**).

### Reduction of outer EECs and metallothionein expression correlates with loss of contractility in SSc

We next evaluated whether the cellular and molecular changes we observed between SSc, GERD, and HCs were correlated with specific clinical measures of esophageal dysfunction (**Table 1**). To more efficiently model the covariance structure among quantitative, functional esophageal measures in SSc (**Figure 5A**), we performed PCA on a set of nine clinical metrics and tested for correlations with the first two PCs (**Figure 5B**). The first PC explained 45.5% of variance and was most correlated with FLIP intra-balloon pressure at 60ml (r=0.94), The second PC explained 22.6% of variance and was most correlated with HRM basal EGJ pressure (r=0.74).

**Figure 5.**
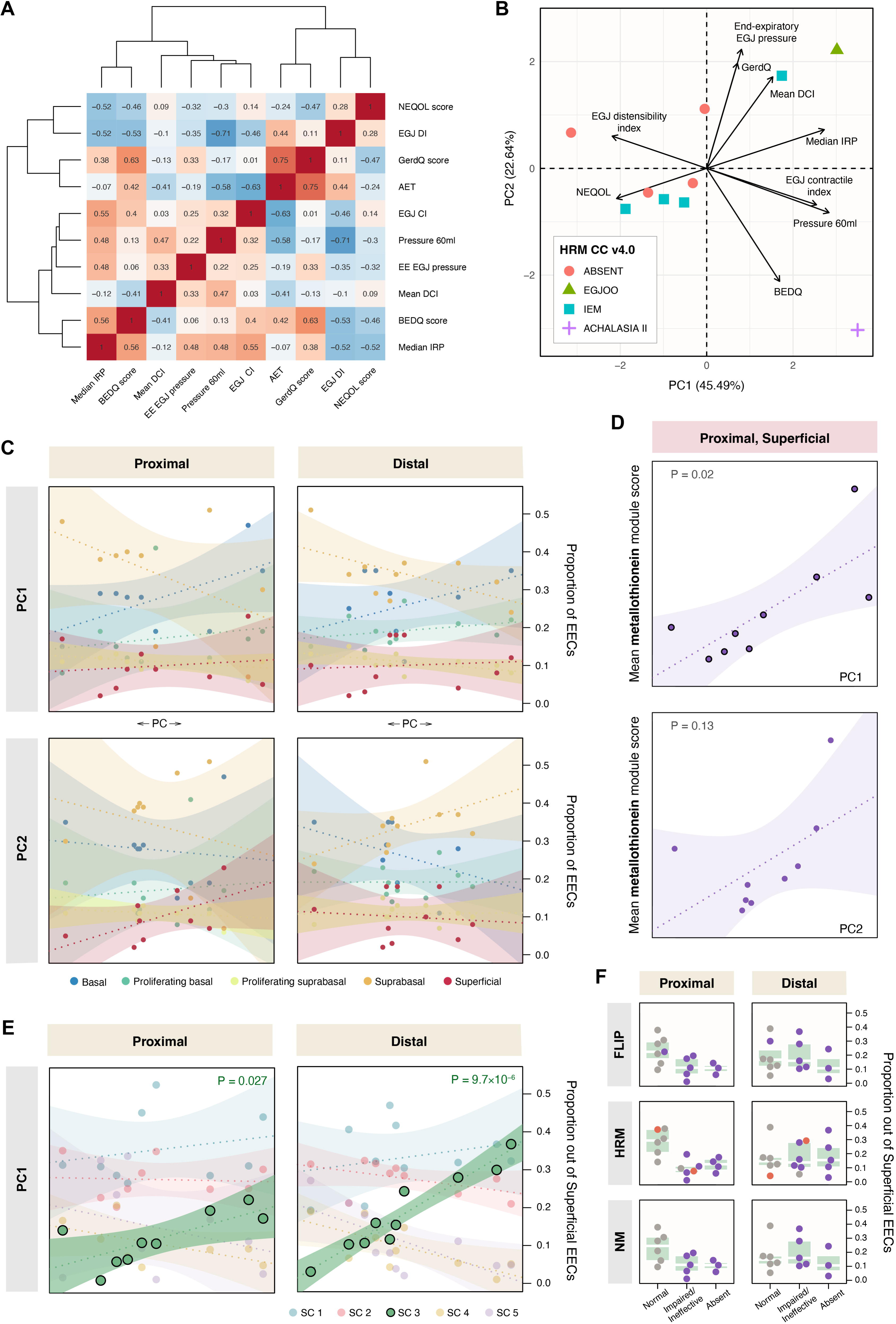
Correlations with clinical phenotypes. **A)** Heatmap of quantitative clinical traits organized using agglomerative, complete hierarchical clustering on Euclidean distances, with pairwise Spearman correlations shown in each cell. **B)** SSc cases are plotted on the first two PCs of the quantitative clinical traits, colored by esophageal motility phenotype. The relative magnitude and direction of trait correlations with the PCs are shown with black arrows. **C)** Correlations between EEC compartments and clinical trait PCs in SSc cases. Linear regression trendlines with 95% confidence intervals are shown for each compartment against PC1 and PC2. **D)** Correlations between aggregate, relative metallothionein expression and clinical trait PCs in superficial cells in the proximal esophagus. Trendlines with 95% confidence intervals are shown with the unadjusted correlation P-value. The metallothionein module score included *MT1A*, *MT1E*, *MT1F*, *MT1G*, *MT1H*, *MT1M*, *MT1X*, and *MT2A*. **E)** Correlations between superficial clusters (SCs) and clinical trait PC1 in SSc cases. Linear regression trendlines with 95% confidence intervals are shown for each SC. The displayed P-values correspond to the highlighted correlations between SC3 and PC1. **F)** Boxplots showing the proportion of SC3 in superficial cells by esophageal motility phenotype in the proximal and distal esophagus, colored by disease state (HCs, grey; GERD, orange; SSc, purple). AET, acid exposure time; BEDQ, brief esophageal dysphagia questionnaire; BMI, body mass index; DCI, distal contractile integral; DI, distensibility index; EGJ, esophagogastric junction; EGJOO, esophagogastric junction outflow obstruction; FLIP, functional luminal imaging probe panometry; HCs, healthy controls; HRM, high-resolution manometry; IEM, ineffective esophageal motility; IQR, inter-quartile range; IRP, integrated relaxation pressure; NEQOL, Northwestern esophageal quality of life; NM, neuromyogenic model. NR, not reported.

We did not observe any significant associations between epithelial cell compartment proportions in SSc and esophageal motility phenotypes. It appeared that the low proportion of superficial cells in the esophageal epithelium was a universal feature in SSc, regardless of phenotype (**Supplemental Figure 7, A-B**). The proportion of superficial cells decreased with disease duration, but the trend was not statistically significant (P=0.1; **Supplemental Figure 7**C). We also did not observe any significant correlations between EEC compartment proportions and quantitative trait PCs (**Figure 5C**).

We then tested for correlations with gene expression in superficial EECs for genes that were significantly differentially expressed in SSc compared to HCs (P_adj_<0.05) and nominally differentially expressed in SSc compared to GERD (P<0.05). There were 433 genes that met these criteria in the proximal esophagus and 99 in the distal esophagus. Although no associations with esophageal motility phenotypes were statistically significant after adjusting for multiple testing, the strongest phenotypic correlation was with *TRIM11* (P_FLIP_=0.01, P_HRM_=0.007, **Supplemental Figure 7D**), a gene recently found to attenuate Treg cell differentiation in CD4^+^ T cells in mice^81^. No associations with the top two clinical PCs were significant after adjusting for multiple testing. We also assessed clinical PC correlations with aggregate metallothionein gene expression and transcription factor target expression for IRF1, MYC, E2F4, and NFE2L2. We observed one nominal association between the mean metallothionein module score and PC1 (P=0.02, **Figure 5D**). Notably, within superficial cells we also observed a strong correlation between the proportion of metallothionein-expressing cluster 3 cells and PC1 in the proximal (P=0.027) and distal esophagus (P=9.7×10^-6^; **Figure 5E**). This indicates that not only is this population of cells reduced in SSc, but its relative decrease is further correlated with more severe esophageal involvement (**Figure 5F**).

## DISCUSSION

Esophageal dysfunction is extremely common in SSc and is significantly associated with increased mortality and lower quality of life^82,83^. The causal mechanisms by which SSc affects the esophagus, however, have not been conclusively determined. Here, we sought to clarify the role of epithelial cells in SSc esophageal dysfunction by using scRNA-seq to quantify cellular and transcriptional changes relative to HCs and individuals with GERD. While our findings suggest that epithelial changes in SSc result primarily from chronic acid exposure, they also highlight immunoregulatory pathways uniquely altered in SSc that may be linked to pathogenic aberrations.

The pathogenic role of epithelial cells in SSc has been an ongoing area of research^23^. When damaged, epithelial cells release signals that help induce fibroblast activation to promote wound healing^84^. In SSc, epidermal keratinocyte characteristics resemble a pro-fibrotic, activated state^85^. SSc epidermal cells were also found to stimulate fibroblasts in culture^86^, and some signs related to epithelial-mesenchymal transition (EMT) have been observed in the SSc epidermis^87^. The FLI1 (friend leukemia integration 1) transcription factor, in particular, has received attention for its potential role in epithelial-cell-mediated SSc pathogenesis^23^. Lower FLI1 expression was observed in the epidermis of diffuse cutaneous SSc, and inactivation of FLI1 in human keratinocytes in vitro induced gene expression changes characteristic of SSc^25^. Furthermore, conditional deletion of *Fli1* in epithelial cells produced an SSc-like phenotype in mice^25^. Importantly, the epithelial-cell-specific *Fli1* knockout further recapitulated the esophageal involvement of SSc, with increased collagen deposition in the lamina propria combined with atrophy of the circular muscle layer, and the esophageal changes were not mediated by T-cell autoimmunity^25^. In contrast to these previous studies in mice and epidermal keratinocytes, we did not observe *FLI1* expression in EECs in any condition. *FLI1* was expressed at low levels in endothelial cells, but the expression differences between SSc, HCs, and GERD were not significant. We did observe significantly increased expression in SSc and GERD of the *PI3* gene, which encodes trappin-2/elafin, previously found to be induced by Fli1 silencing in human dermal microvascular endothelial cells^88^. However, this association is unlikely to be pathogenic, as the increase in gene expression was greater in GERD, and a growing body of evidence indicates that trappin-2/elafin is expressed to promote tissue repair in response to gastrointestinal tissue inflammation^89^.

While there have been many studies of epithelial cells in skin in SSc, molecular study of the human esophagus in SSc has heretofore been limited to one array-based gene expression study in bulk tissue conducted by Taroni and colleagues^15^, in which they identified distinct expression signatures among variable genes between 15 individuals with SSc that were similar to those seen in skin^90^. These signatures were independent of clinically defined SSc subsets^15^, suggestive of inter-individual heterogeneity in SSc. They also found that gene expression patterns between upper and lower esophageal biopsies within individuals were nearly identical. They did not observe statistical associations between inflammatory gene expression signatures and GERD, but the study was underpowered for such an analysis, and GERD was defined based on histological evidence for basal cell hyperplasia and intraepithelial lymphocyte counts.

The gene expression differences we observed in SSc epithelial cells were highly correlated with those seen in GERD, and nearly all differential gene expression was limited to the superficial layers of the epithelium. These observations suggest that the primary driver of differential gene expression in EECs in SSc is chronic acid exposure. The genes with the strongest mutual upregulation in SSc and GERD were primarily related to keratinization/cornification, including small proline-rich proteins (SPRRs), serine protease inhibitors (serpins), S100 proteins, late cornified envelope (LCE) genes, and keratins associated with terminal epithelial differentiation, namely *KRTDAP* and *KRT*. *KRT1* helps maintain epithelial barrier function in gastrointestinal epithelial cells, and its overexpression was shown to attenuate IL-1β-induced epithelial permeability^91^. The mutual upregulation of these genes in the outer layers of the epithelium therefore likely represent a protective response to chronic acid exposure.

While the overall gene expression differences were highly correlated between SSc and GERD, there were sets of genes disproportionately upregulated in SSc that hint at disease-related immune response aberrations, including genes that have previously been implicated in SSc pathogenesis. For example, *PTGES*, which encodes prostaglandin E synthase, was previously found to be significantly upregulated in SSc fibroblasts^92^ and inflammatory non-classical monocytes^93^, and Ptges-null mice were resistant to bleomycin-induced fibrosis^92^. CD44 and CD74 form the receptor complex for the macrophage migration inhibitory factor (MIF), an inflammatory cytokine that promotes fibroblast migration and has been implicated in SSc pathogenesis^94^. LTB4R (Leukotriene B4 Receptor, or BLT1) has been found to activate AKT/mTOR signaling and knockdown of *LTB4R* attenuated fibrosis in bleomycin-induced SSc in murine models^95^. Serum HBEGF (heparin-binding epidermal growth factor) levels and fibroblast *HBEGF* expression were both significantly elevated in SSc^96^. Copy number variation of *APOBEC3A* was significantly associated with SSc in a Han Chinese population^97^. Finally, the SSc-specific upregulation of genes involved in antigen processing and presentation (*HLA-B*, *CD74*, *TAP1*, *PSMB8*, *PSMB9*) are suggestive of increased human leukocyte antigen (HLA) class I activity in superficial EECs in SSc. Notably, *TAP1*, *PSMB8*, and *PSMB9* are located immediately adjacent to one another in the class II region of the HLA locus, which is the genomic region with the strongest genetic associations with SSc^98^, and SSc-associated HLA-B alleles have also been identified^99,100^.

The most striking set of genes that were uniquely downregulated in SSc were metallothioneins. The expression of detected metallothioneins (*MT1A*, *MT1E*, *MT1F*, *MT1G*, *MT1H*, *MT1M*, *MT1X*, *MT2A*) was strongly reduced in the superficial compartment of EECs in the proximal esophagus in SSc compared to HCs. We further observed a nominal association between relative metallothionein expression in the proximal esophagus and the first PC of esophageal dysmotility among SSc patients, with lower expression of metallothioneins correlated with greater EGJ distensibility and weaker contractility. The proportion of the primary metallothionein-expressing cluster within superficial EECs was likewise significantly associated with clinical PC1. The metallothioneins are a family of well-conserved, metal-binding proteins that regulate zinc and copper homeostasis, prevent heavy metal poisoning, and combat oxidative stress^101,102^. Their study in a wide array of physiological conditions and immune-mediated diseases has revealed the proteins play central roles in innate and adaptive immunity, including autoimmunity, but their immunoregulatory behavior is highly complex and context-specific^101,102^. Given their import in immune regulation, their associations with SSc and esophageal motility reported in this study, and the connection between SSc and heavy metal exposure^103^, the metallothioneins are compelling candidates that warrant further study in SSc.

Interestingly, there was greater relative gene dysregulation in the proximal esophagus compared to the distal esophagus in SSc, but the opposite was true for GERD. This discrepancy suggests that esophageal dysmotility in SSc leads to greater acid exposure in the proximal esophagus, since refluxate typically affects the distal esophagus significantly more than the proximal esophagus^104^ and abnormal peristalsis prolongs acid clearance^105^. This finding coincides with a bulk RNA-seq study of esophageal mucosa of achalasia patients^106^, which likewise identified more differential expression in the proximal esophagus than in the distal esophagus, including significant enrichment of matrisome-associated genes and lower expression of genes associated with reactive oxygen species metabolism^106^. Due to differences in innervation depth^107^, the epithelium of the proximal esophagus has more nociceptive sensitivity than the distal esophagus, which likely contributes to the association between gastrointestinal symptoms and lower health-related quality of life in SSc^108^. Proximal acid exposure increases the risk of aspiration^109^ and is significantly more prevalent in SSc patients with idiopathic lung fibrosis^110^.

Targets for several transcription factors, including IRF1, MYC, E2F4, and NFE2L2 (or NRF2), were significantly enriched among DEGs in superficial EECs from the proximal esophagus. IRF1 is involved in innate immune responses, but while IRF1 targets were enriched in DEGs between SSc and GERD, the enrichment appeared to be driven by upregulation in GERD. IRF1 targets were not enriched in DEGs between SSc and HCs, nor was *IRF1* significantly differentially expressed in SSc. MYC, E2F4, and NFE2L2 all participate in different stages of the cell cycle^111–113^, and expression of each was highest in either the proliferating basal or suprabasal compartments, but these proteins may also play a role in terminal differentiation of EECs, particularly under conditions of stress. In the least differentiated superficial cluster (cluster 5), where the expression differences relative to HCs for each of these transcription factors and their targets was most pronounced, the expression level changes in GERD were correlated with those observed in SSc. Furthermore, Myc ablation in Krt14-expressing tissues in mice led to abnormal terminal differentiation of epithelial cells^114^, and studies of the esophageal epithelium in Nrf2-knockout mice indicate that Nrf2 regulates cornification of keratinized epithelial cells under oxidative stress^115,116^. Moreover, *C10orf99*—one of the genes uniquely upregulated in SSc with superficial expression mirroring *MYC*, *E2F4*, and *NFE2L2*—was found to increase expression of proinflammatory markers and attenuate late differentiation of keratinocytes under conditions of stress^117^. These findings therefore suggest that within the SSc esophagus there is activation of pathways that regulate the late differentiation of EECs in response to external stress, and these could contribute to the reduction of superficial EECs we observed in SSc.

There are important limitations to consider when interpreting this study. First, this study only focused on squamous epithelial cells. There may be relevant changes in other cell types within the esophageal mucosa that correlate with SSc and dysmotility, but such analyses are ongoing and were outside the scope of this investigation. Second, the relatively small patient sample size of this study combined with the heterogeneity of SSc subtypes and dysmotility phenotypes limited our power to detect clinical associations between cell type proportions or gene expression changes and specific clinical phenotypes. Heterogeneous molecular profiles in SSc esophagus samples have been described previously^15^, but we grouped all SSc patients together in our analyses for comparisons against GERD and HCs. We applied ordinal logistic regression on motility classifications and performed principal components analysis on reflux- and motility-related quantitative traits to increase our power to detect clinical associations but were nonetheless statistically limited by sample size. Third, our analysis of imaging data in the SSc esophageal mucosa was limited. While the spatial molecular imaging confirmed our cell annotations and reproduced our findings within the superficial compartment, CosMx’s cell segmentation algorithm could not aptly capture the flattened, densely packed outer cell layers of the epithelium, resulting in a relatively sparse representation of superficial EECs. Furthermore, the spatial analysis was limited to 8 samples from 4 individuals. Therefore, we cannot make more robust conclusions on the morphological states that accompany the transcriptional and cellular proportion changes that we observed. Future studies utilizing larger, more homogeneous cohorts with complete imaging data may be able to detect more esophageal epithelial changes associated with dysmotility in SSc.

In summary, esophageal complications are common in individuals with SSc and can greatly impact quality of life and lifespan. In this study, we sought to deepen our understanding of the role of the epithelium in SSc esophageal involvement. Through a thorough, single-cell-level transcriptomic investigation, we identified the unique cellular and transcriptional differences present in the SSc esophageal epithelium. There were significantly fewer terminally differentiated epithelial cells in the apical, superficial layers in both the proximal and distal esophagus, but otherwise epithelial compartment proportions were similar to healthy controls, including proliferating cell proportions. Significant differences in gene expression were likewise almost exclusively found in the superficial compartment, and these changes were highly correlated with those seen in individuals with GERD, indicating that the primary driver of differential gene expression in the SSc esophageal epithelium is chronic acid exposure. Transcriptional differences were more prominent in the proximal esophagus of SSc patients than in GERD patients, possibly due to greater proximal exposure resulting from esophageal dysmotility. Genes that were most disproportionately dysregulated in SSc compared to GERD belonged to immune-related pathways, including innate and adaptive response, antigen presentation, and metal homeostasis, pointing to pathogenic immune dysfunction. Future studies investigating these pathways in non-epithelial cell populations in more homogeneous SSc cohorts may further resolve the pathogenesis of esophageal involvement in SSc. By serving as an atlas for the human esophageal epithelium in SSc, this study can guide future efforts to address remaining gaps in our understanding of the SSc esophagus, to ultimately uncover pathogenic mechanisms and identify actionable targets.

## Data Availability

Raw and processed sequencing data with corresponding metadata will be deposited in a genomic data repository upon study publication in a peer-reviewed journal.

---

**Supplemental Figure 1.**
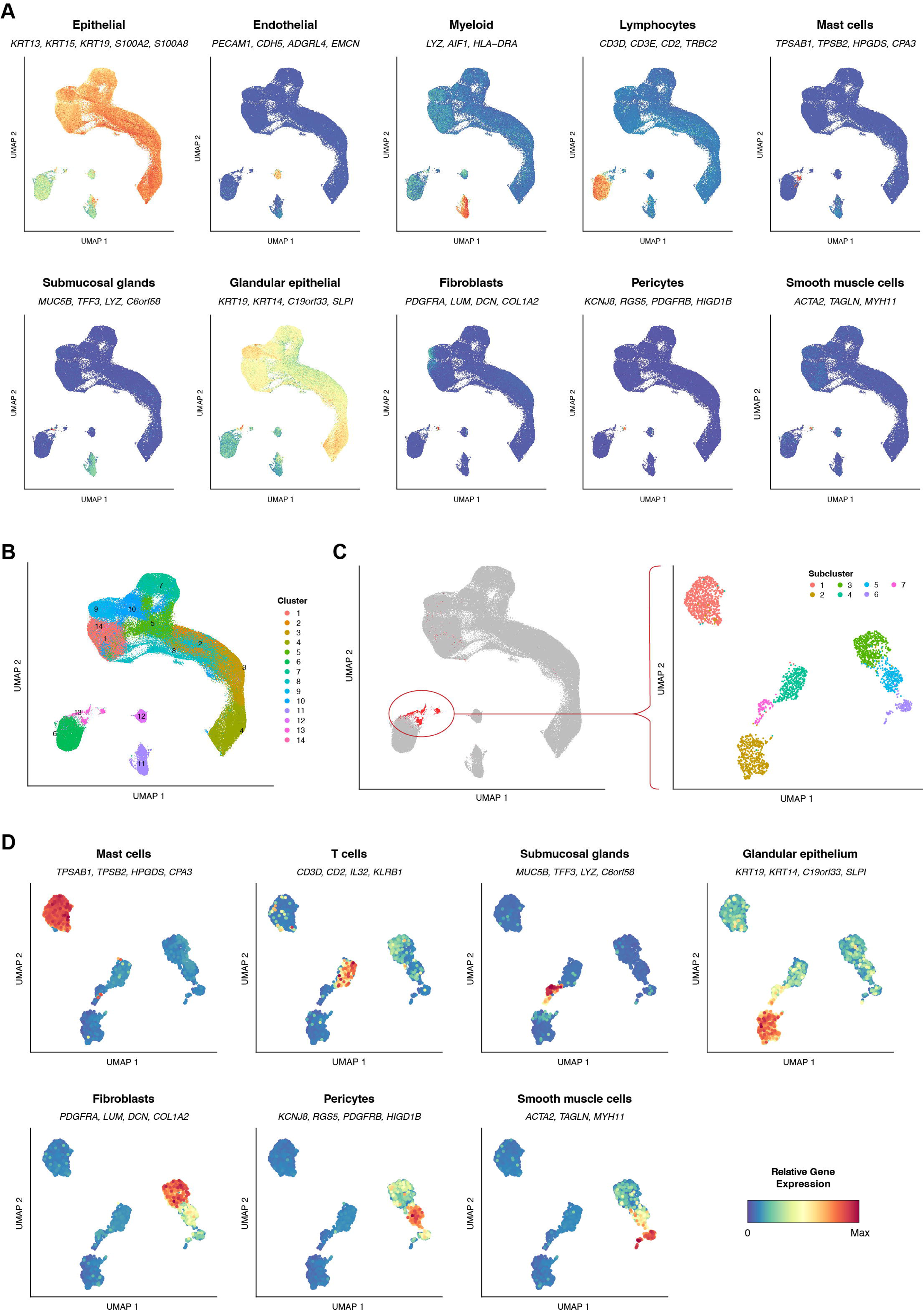
Cell clustering and cell type annotation. **A)** Cell type module scores for 10 detected cell types, colored by relative gene expression of the listed genes. **B)** Projected UMAP embeddings calculated on the top 40 PCs with n=200 neighbors. Cells are colored by cluster, which are numbered in descending order by total cell count. Clusters were determined using Seurat’s modularity-based clustering on a shared nearest neighbor (SNN) graph with k=25 and resolution=0.25 on the top 40 PCs. **C)** Isolation of cluster 13 for annotation of cell types with smaller proportions of cells and projected UMAP embeddings for cluster 13, colored by subcluster. **D)** Cell type module scores for 7 detected cell types in subcluster 13, colored by relative gene expression of the listed genes.

**Supplemental Figure 2.**
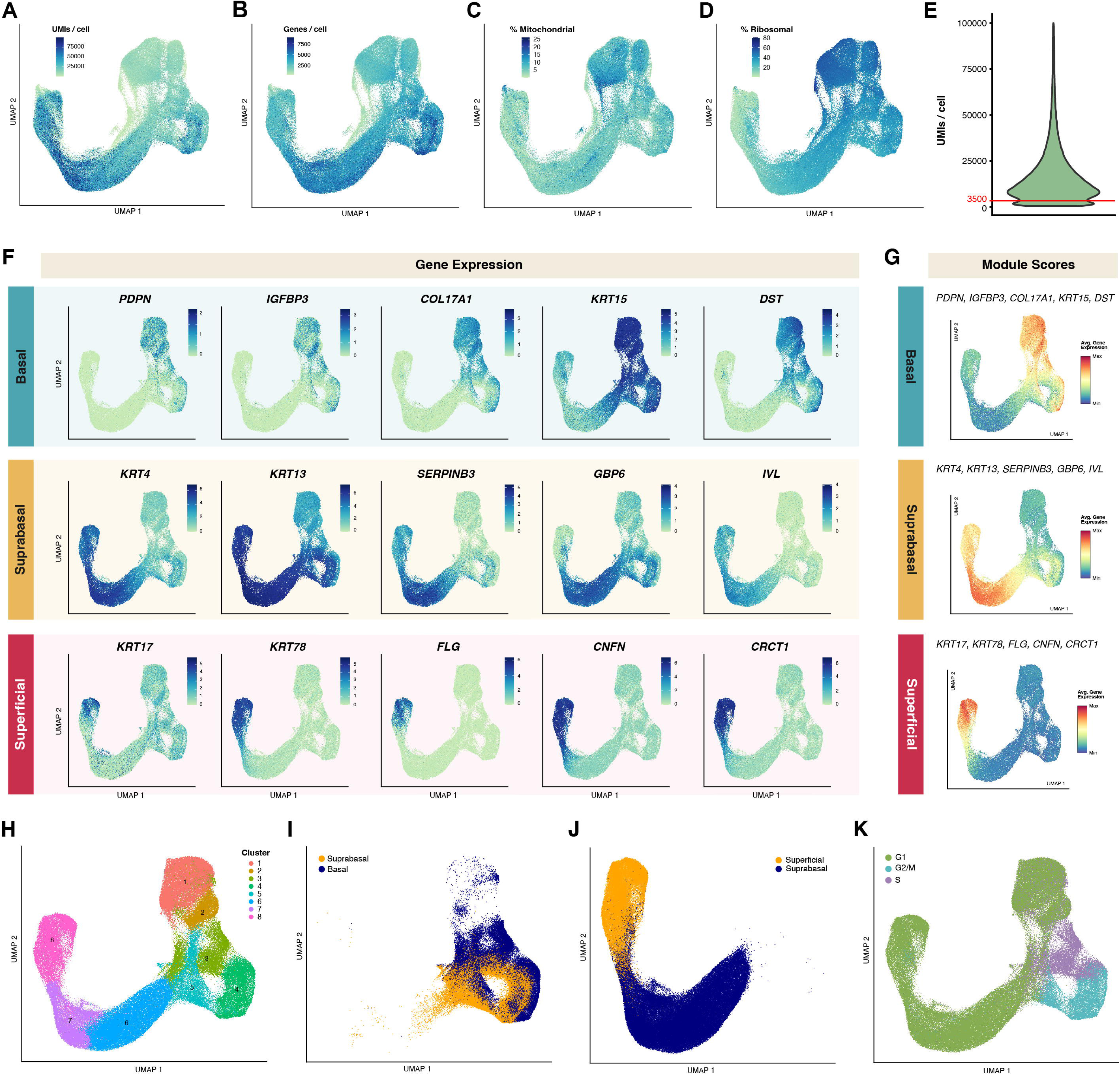
Epithelial cell quality control and annotation. **A)** EEC UMAP projection, colored by the number of unique molecular identifiers (UMIs) per cell. **B)** EEC UMAP projection, colored by the number of detected genes per cell. **C)** EEC UMAP projection, colored by the number of proportion of UMIs mapping to mitochondrial DNA. **D)** EEC UMAP projection, colored by the proportion of UMIs mapping to ribosomal DNA. **E)** Distribution of number of UMIs detected per cell. EECs with <3500 UMIs/cell were removed from further analysis. **F)** EEC UMAPs colored by gene expression (log(counts+1))) for genes included in compartment-specific expression scores. **G)** EEC UMAPs colored by compartment expression scores. **H)** Projected UMAP embeddings for post-quality-control EECs calculated on the top 35 PCs with n=100 neighbors. Cells are colored by cluster, which are numbered in descending order by total cell count. Clusters were determined using Seurat’s modularity-based clustering on a SNN graph with k=50 and resolution=0.25 on the top 35 PCs. **I)** Result of K-means clustering to distinguish suprabasal from basal cells, based on expression of corresponding cluster markers (**F**) in clusters 3, 4, and 5, with K=2. **J)** Result of K-means clustering to distinguish superficial from suprabasal cells, based on expression of corresponding cluster markers (**F**) in clusters 6, 7, and 8, with K=2. **K)** Predicted classification of cell cycle state in EECs determined using Seurat’s CellCycleScoring() function.

**Supplemental Figure 3.**
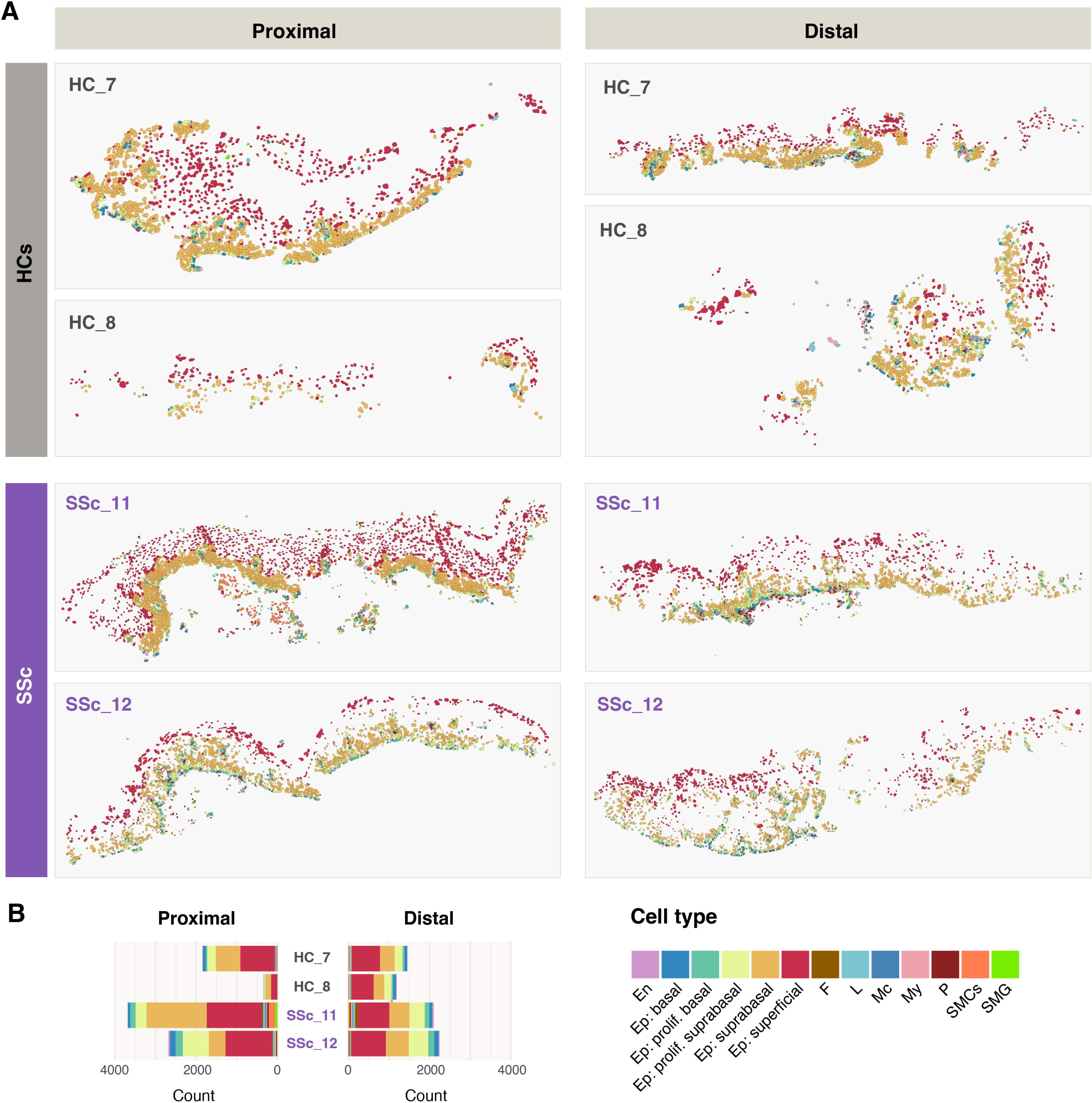
Spatial molecular imaging of esophageal mucosa. **A)** Esophageal mucosa tissue slides analyzed by CosMx spatial molecular imaging are shown with annotations conferred from scRNA-seq label transfer for two healthy controls (HCs) and two SSc patients with absent contractility **B)** Spatial molecular imaging cell type counts are shown by individual and biopsy location. En, endothelial; Ep, epithelial; F, fibroblast; L, lymphocytes; Mc, mast cells; My, myeloid; P, pericytes; SMCs, smooth muscle cells; SMG, submucosal glands.

**Supplemental Figure 4.**
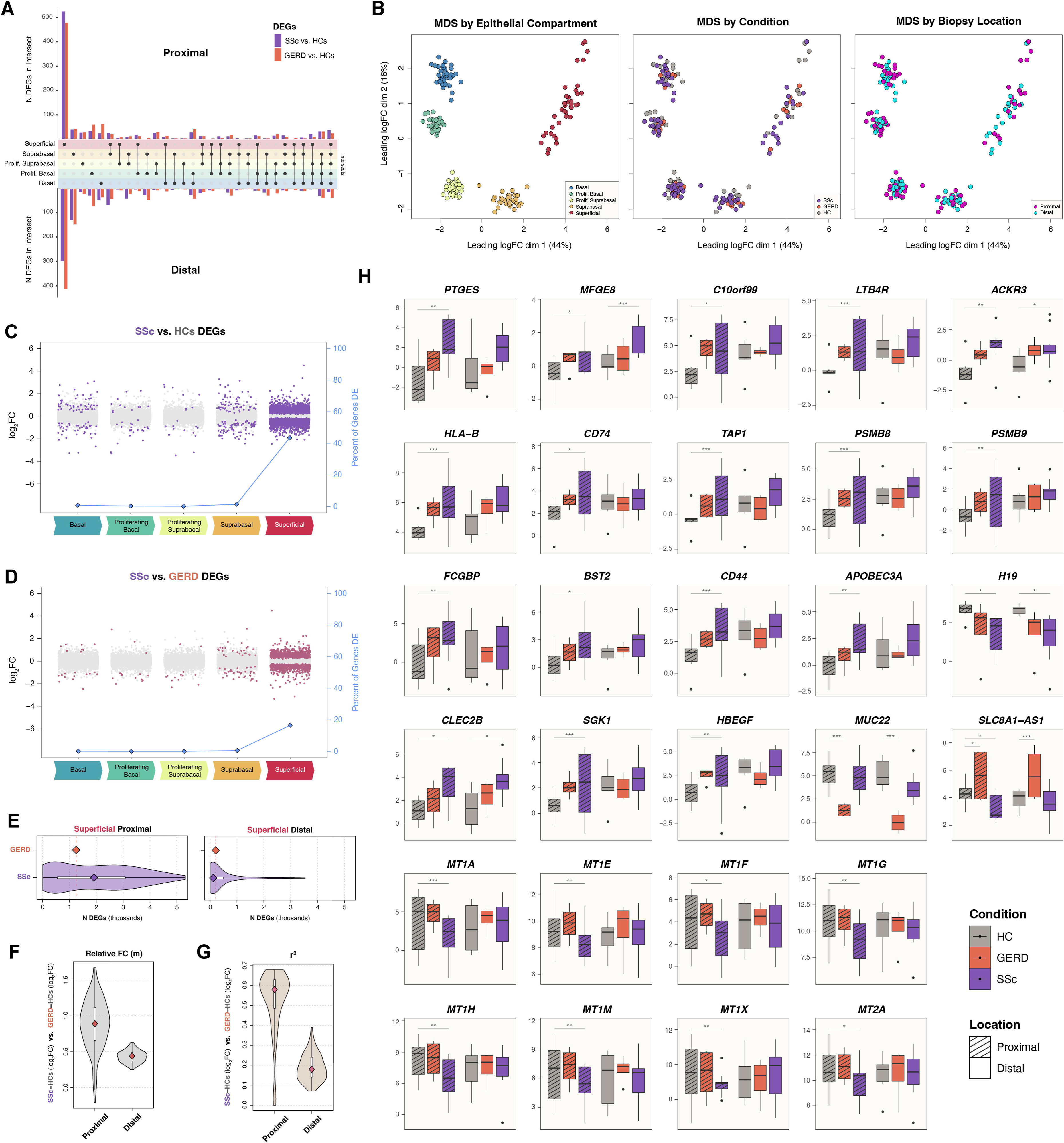
Group-wise gene expression correlation for variable genes. **A)** UpSet plots showing differential gene expression at the single-cell level by epithelial compartment for SSc vs HCs (purple) and GERD vs HCs (orange) in the proximal and distal esophagus. **B)** Multidimensional scaling plots of aggregate gene expression samples colored by sample, epithelial compartment, condition, and biopsy location. **C-D)** Pseudobulk gene expression differences between SSc and HCs **(C)** and between SSc and GERD **(D)** are shown for each EEC compartment. Significant DEGs (FDR<0.05) are highlighted in purple and raspberry, respectively. The percentages of genes that were significantly differentially expressed in each compartment are plotted in blue in along the secondary Y axis. **E)** The number of significant DEGs are shown for GERD vs. HCs (orange) and for all permutations of n=4 samples for SSc vs. HCs (purple) in superficial cells for both proximal and distal regions. The point within the SSc distribution is the median value. **F-G)** The distributions of relative FC or slope *m* **(F)** and coefficient of determination *r^2^* **(G)** are shown for all permutations of n=4 SSc samples, determined by modeling a linear regression with intercept=0 for the log_2_FC of SSc vs. HCs against the log_2_FC of GERD vs. HCs for all expressed genes in the superficial compartment for both proximal and distal regions. DEG, differentially expressed gene; FC, fold change. **H)** Boxplots of pseudobulk gene expression, aggregated by condition, biopsy location, and EEC compartment for disease-specific DEGs referenced in the main text. Pairwise differential expression was determined using edgeR’s quasi-likelihood F-tests: *p<0.05; **p<0.05; ***p<0.0005.

**Supplemental Figure 5.**
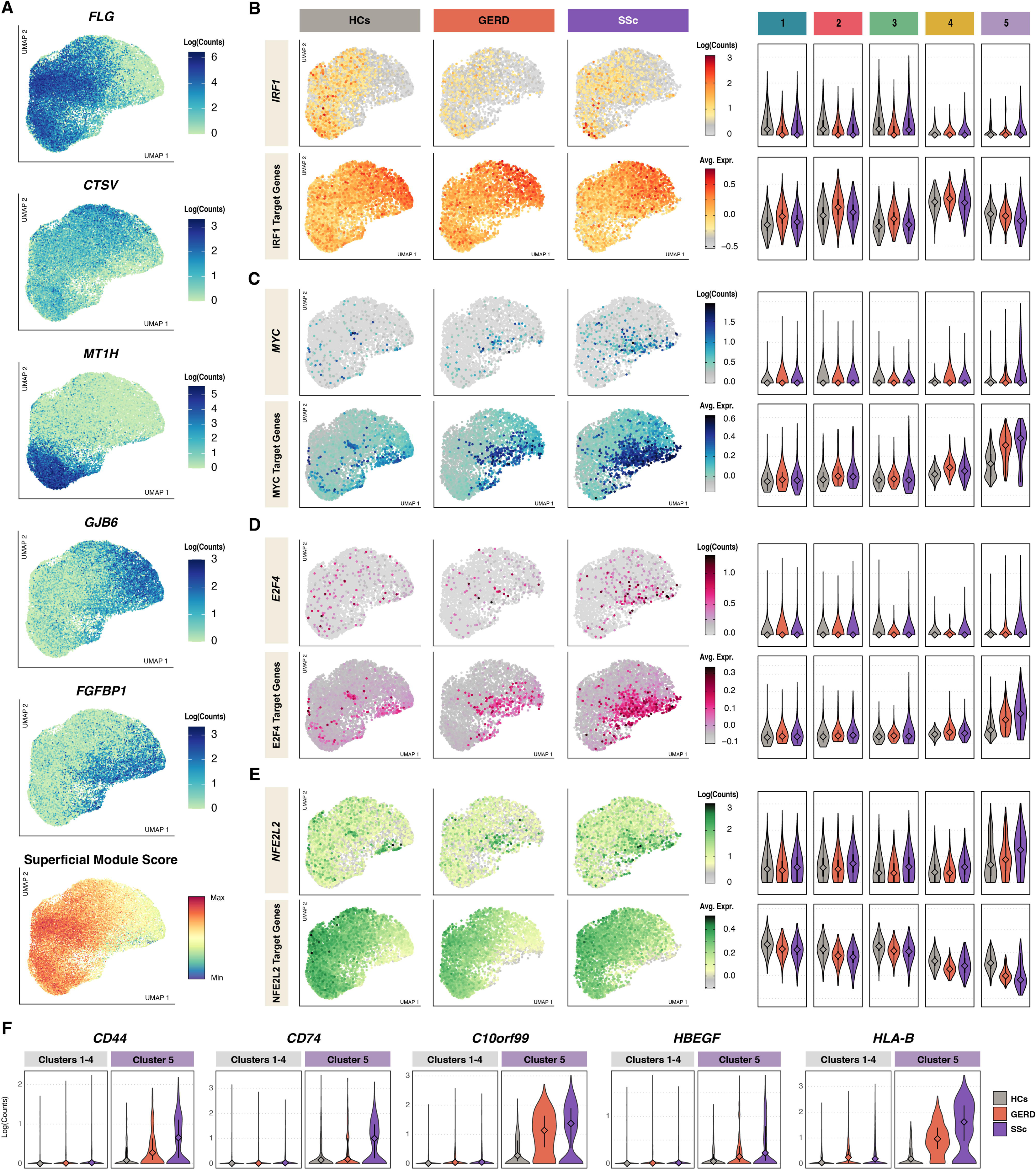
Expression of cluster markers, enriched transcription factors, and their targets in superficial EECs in proximal esophagus. **A)** UMAPs with expression for the top superficial cluster markers are shown along with the epithelial differentiation score for superficial cells. **B-D)** The distributions of enriched transcription factor expression and target module scores are shown proximal, superficial cells by condition and superficial cluster for **B)** *IRF1*, **C)** *MYC*, **D)** *E2F4*, and **E)** *NE2L2*. **F)** Violin plots of gene expression in superficial cluster 5 vs. all other superficial subclusters for SSc-specific differentially expressed genes that were uniquely expressed in cluster 5 within superficial EECs.

**Supplemental Figure 6.**
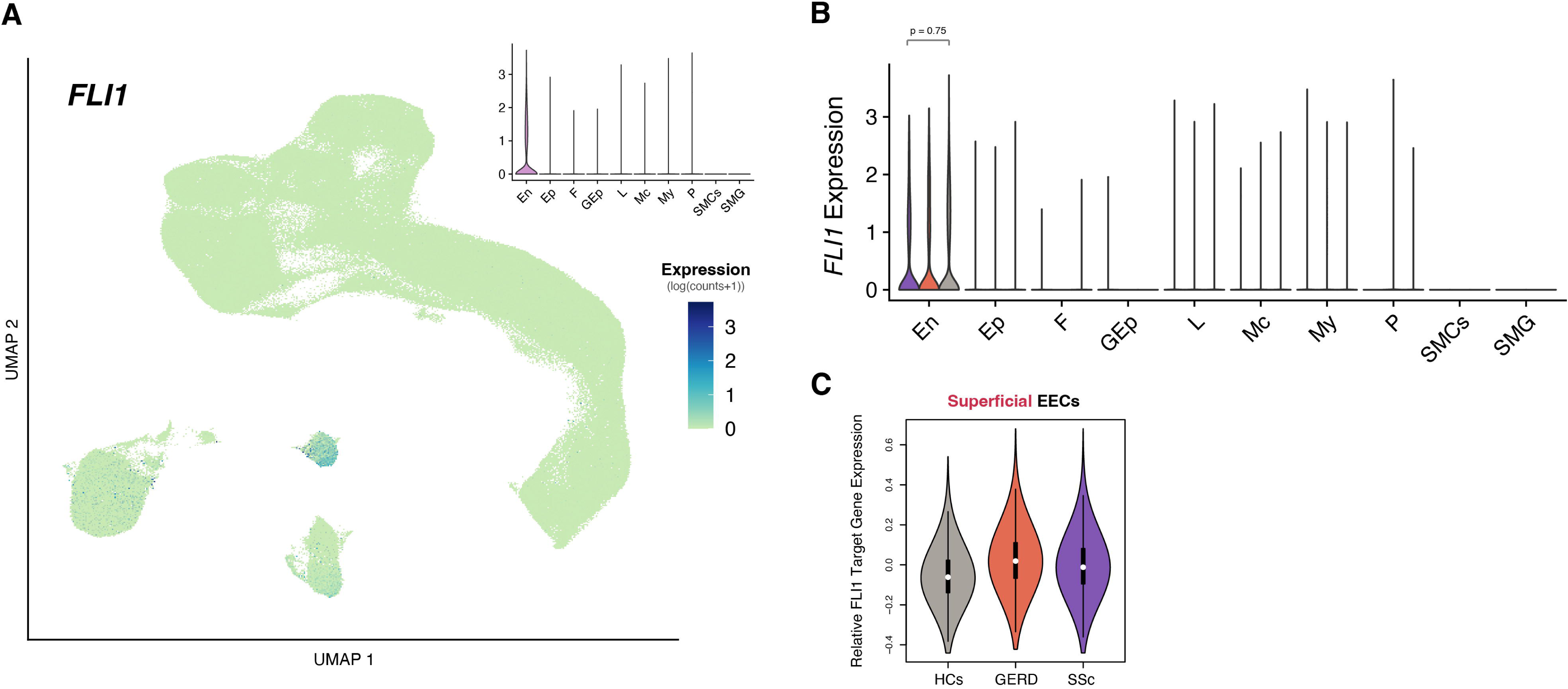
*FLI1* expression in human esophageal mucosa. **A)** UMAP of integrated mucosal dataset, colored by *FLI1* expression. The distribution of *FLI1* expression by cell type is shown in the upper right. **B)** Distribution of *FLI1* expression by cell type and disease state. Endothelial cells had the highest *FLI1* expression, but the expression differences between diseases was not significant. **C)** Violin plot of the relative expression of FLI1 target genes in superficial cells.

**Supplemental Figure 7.**
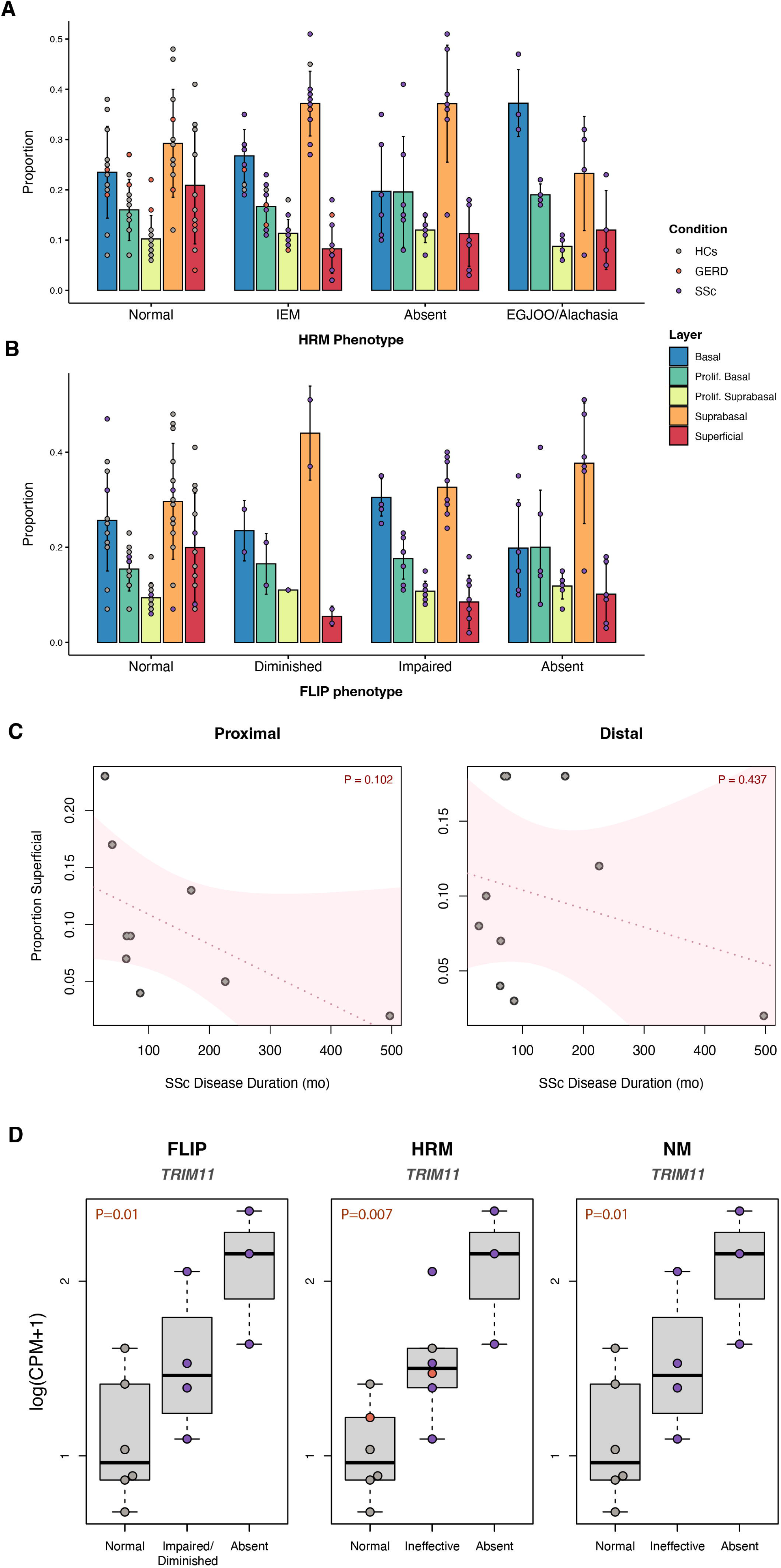
Epithelial cell compartment proportions and additional correlations by motility phenotype. **A-B)** Proportions of esophageal epithelial cell compartments by HRM **(A)** and FLIP **(B)** esophageal dysmotility phenotypes are shown for the proximal and distal esophagus samples. The bars denote the mean values, the vertical lines the standard deviations, and the points the individual sample proportions. **C)** The relationship between the proportion of superficial cells and the SSc disease duration is plotted for proximal and distal samples. Trendlines with 95% confidence intervals are plotted in pink, and correlation P-values are displayed in the upper right corners. **D)** The distribution of normalized gene expression values by esophageal motility phenotype are shown for the *TRIM11* gene for the FLIP, HRM, and NM esophageal dysmotility classifications. Unadjusted ordinal regression p-values are displayed in the upper left corners. *TRIM11* was the most strongly association gene with the dysmotility phenotypes by p-value. HRM, high-resolution manometry; FLIP, functional luminal imaging probe panometry; IEM, ineffective esophageal motility; EGJOO, esophagogastric junction outflow obstruction; NM, neuromyogenic model.

